# A Distinct Form of Subcutaneous Fat Fibrosis Predicts Insulin Resistance in People with HIV

**DOI:** 10.1101/2025.05.13.25327547

**Authors:** Diana L. Alba, Moon K. Choi, Alaa Abdellatif, Stephen Brown Mayfield, Thuy An T. Pham, David Berrios, Antonio Rodriguez, Marin Ewing, Tony Figueroa, Judy Gonzalez-Vargas, Ningyan Zhang, Zhiqiang An, Dawei Bu, Steven G. Deeks, Philipp E. Scherer, Peter W. Hunt, Suneil K. Koliwad

## Abstract

**Background:** People with HIV (PWH) are at heightened risk for type 2 diabetes (T2D) and insulin resistance (IR), even with effective antiretroviral therapy (ART). Adipose tissue dysfunction, including subcutaneous adipose tissue (SAT) fibrosis, is a key contributor to metabolic disease. However, the role of SAT fibrosis in IR among PWH remains poorly understood. We therefore investigated the relationship between SAT fibrosis and IR in PWH along with molecular signatures that might distinguish HIV-associated SAT fibrosis from that associated with obesity.

**Methods:** We studied 112 participants, including 43 PWH and 69 people without HIV (PWoH) and excluding those with established T2D. Body composition was assessed by dual-energy X-ray absorptiometry (DXA), and SAT fibrosis was analyzed by quantifying hydroxyproline levels from biopsies. SAT transcriptional profiles were examined using a targeted fibrosis-related gene panel. Plasma levels of endotrophin, a marker of extracellular matrix remodeling, were also measured.

**Results:** PWH had significantly greater SAT fibrosis compared to PWoH, with the largest differences observed among participants without obesity. In this subgroup, SAT fibrosis was strongly associated with IR, despite the absence of elevated adiposity. Transcriptomic analysis identified a distinct fibrosis-associated gene expression pattern in PWH, marked by upregulation of *COL14A1* and key immune-related genes (e.g. *CCL4*, *NLRP3*). Pathway analysis further supported upregulated extracellular matrix remodeling and immune activation, along with downregulated thermogenesis, lipid metabolism, and insulin signaling in the SAT of non-obese PWH. Plasma endotrophin levels were significantly elevated in PWH and were independently associated with SAT fibrosis.

**Conclusion:** Our findings identify SAT fibrosis as an obesity-independent driver of IR in PWH. Notably, SAT fibrosis predicts IR at normal body fat levels and can be noninvasively monitored through circulating endotrophin, offering a potential biomarker for early intervention. The distinct transcriptional signature of HIV-associated fibrosis reveals unique mechanisms that may underlie heightened metabolic risk in this population and highlight new therapeutic avenues targeting adipose tissue remodeling and metabolic dysfunction.

## Introduction

People living with human immunodeficiency virus (HIV) infection exhibit higher rates of metabolic diseases, including cardiovascular disease, hypertension, and type 2 diabetes mellitus (T2D), compared to people without HIV (PWoH)(1, 2). Notably, this increased risk is not confined to untreated HIV infection but is also seen in people achieving durable viral suppression through effective antiretroviral therapy (ART)(3, 4). Several studies have reported that people living with ART-controlled HIV face a 2- to 4-fold higher risk of incident T2D than PWoH(1, 3, 5) emphasizing that metabolic complications are a significant concern for PWH despite effective viral suppression.

Beyond the well-established drivers of insulin resistance (IR) and T2D in the general population, additional unique factors amplify disease risk for PWH. One such factor is the use of particular ART regimens. For example, certain protease inhibitors, such as indinavir, lopinavir, and ritonavir, have been shown to reversibly induce insulin resistance by inhibiting glucose transport via GLUT4(6–8). This inhibition disrupts normal glucose uptake, contributing directly to metabolic dysfunction in PWH who take these forms of ART(9, 10). Nucleoside reverse transcriptase inhibitors (NRTIs), such as stavudine (D4T) and zidovudine (AZT), have been implicated, both directly and indirectly, in disrupting glucose metabolism(11, 12). These early-generation NRTIs notably promoted the loss of adipose tissue (AT) mass within the subcutaneous compartment (SAT), with a compensatory increase in visceral adipose tissue (VAT) mass, a redistribution of bodily fat stores that independently elevates the risk of T2D(4). This fat redistribution, termed HIV-associated lipodystrophy, underscored the role of ART in driving metabolic complications in people with HIV. Components of modern ART regimens continue to be examined for potential metabolic effects. Integrase-strand-transfer-inhibitors (INSTIs), for example, were shown to blunt the capacity of adipocyte progenitors to differentiate into metabolically healthy, thermogenic “beige” adipocytes(13) and have been associated with SAT fibrosis in both PWH and non-human primate models of HIV(14). While INSTI-based ART has also been linked to a higher risk of T2D than comparator regimens in large cohorts, there remains disagreement across studies as to whether any such effects are independent of differences in BMI(15, 16).

Importantly, however, the prevalence of T2D among PWH has risen progressively despite substantial shifts in ART composition over the past few decades, underscoring the potential role of HIV itself in driving metabolic dysfunction. Specifically, there is growing recognition that HIV infection directly affects the metabolic function and health of AT. Recent studies have identified ATs as potential reservoirs for HIV, and shifts in the immune cell composition of ATs have been documented in PWH(17, 18). These findings have led to the emerging concept that AT dysfunction in PWH encompasses not only the effects of ART, but also includes complex HIV-specific influences on AT physiology and inflammation that may be distinct from what is experienced by PWoH in response to worsening obesity.

Notably, AT dysfunction is an established driver of IR and T2D in the general population(19–21). Obesity induces AT remodeling, a process that includes adipocyte hypertrophy, immune cell infiltration, fibrosis, and hypoxia, each of which is associated with the development of IR. For example, enlarged adipocytes release increased levels of free fatty acids and pro-inflammatory cytokines, such as tumor necrosis factor-alpha (TNF) and interleukin-6 (IL-6), which are implicated in disrupting insulin signaling and contributing to systemic inflammation(20, 22). Obesity-associated extracellular matrix (ECM) accumulation results in AT fibrosis—a pathological process associated with both inflammation and impaired metabolic function(23). Excessive accumulation of ECM components and consequent AT fibrosis is postulated to drive AT dysfunction by reducing its structural plasticity and metabolic flexibility. For example, in PwoH, SAT fibrosis has been correlated with the expansion of VAT mass, which is correlated with IR and is inversely associated with the capacity to lose fat mass in response to bariatric surgery(24–28). At the cellular level, organellar dysfunction in adipocytes—such as endoplasmic reticulum and mitochondrial stress—further amplifies these disruptions, highlighting the concept that obesity can collectively impact AT metabolic health across multiple anatomical scales(29–31). Whereas systemic inflammation has been shown to persist in PWH despite effective viral suppression(32), the role of SAT fibrosis in the metabolic health of PWH remains poorly understood. By focusing on this gap in knowledge, we may uncover potential therapeutic targets to mitigate rising T2D rates among PWH.

In this study, we therefore leverage a powerful multiethnic cohort of PWH and uninfected controls individuals spanning across a wide range of adiposity allowing us to comprehensively analyze the level of SAT fibrosis in PWH and examine its role in IR. Using transcriptomic analyses, we identify molecular pathways in the SAT that are linked to IR in PWH even in the absence of obesity, excluding individuals with established T2D from our analysis, we are able to focus on early drivers of metabolic dysfunction, isolating the effects of HIV and ART away from responses to obesity or T2D per se. Our findings highlight SAT fibrosis as a key contributor to metabolic risk in PWH, with potential implications for targeted interventions to reduce T2D burden in this high-risk group.

## Results

### Participant characteristics

We studied 112 participants, including 69 PWoH and 43 PWH, as summarized in Table 1. All those in the PWH group had maintained plasma HIV RNA levels below the detectable threshold for a minimum duration of one year before enrollment. The two groups were well-matched for key clinical parameters related to glucose homeostasis, including fasting glucose and insulin levels, Homeostatic Model Assessment for Insulin Resistance (HOMA-IR), and hemoglobin A1c (HbA1c), reflecting our sampling strategy. Similarly, the two groups had a comparable mean BMI and total body fat percentage (%BF), measured using dual-energy x-ray absorptiometry (DXA). Notably, the PWH group was older on average than the PWoH group (p < 0.01) and had a higher proportion of males (p 0.02). The racial composition between our study cohorts were also different, with a greater proportion of Asian participants in the PWoH group, vs a higher percentage of Black participants in the PWH group (p < 0.01). However, these demographic differences simply mirrored the makeup of the greater population of PWH living in the San Francisco Bay area, including the fact that Black people are impacted by HIV at higher rates than other races(33). During the recruitment phase, strategic efforts were implemented to minimize racial and ethnic variation between the two groups. Apart from these demographic differences, The PWH group exhibited higher central obesity, as indicated by a greater proportion of VAT mass relative to body weight (%VAT mass) and a higher android-to-gynoid fat mass ratio. However, multivariate analysis showed that this increased %VAT mass was driven by advanced age and not by HIV status itself [p < 0.001 for age, p = 0.88 for HIV status (Supplemental Table 1)]. These findings suggest that HIV infection does not independently promote excess visceral adiposity in PWH, but rather that age-related fat redistribution impacts PWH, including those in our analysis, as it does in other populations.

**Table 1.** Characteristics of Study Participants.

| *Parameter* | *PWoH (n=69)* | *PWH (n=43)* | *p-value* |
| --- | --- | --- | --- |
| **Age (years)** | 46 ± 13 | 54 ± 10 | <0.01 |
| **Sex at birth (Female/Male)** | 33/36 | 10/33 | 0.02 |
| **Race** |  |  | <0.01 |
| Asian | 26 | 1 |  |
| Black | 3 | 16 |  |
| White | 34 | 18 |  |
| More than one race | 5 | 4 |  |
| Other | 1 | 4 |  |
| **Ethnicity** |  |  | 0.51 |
| Hispanic or Latino | 11 | 8 |  |
| Not Hispanic or Latino | 56 | 35 |  |
| Unknow | 2 | 0 |  |
| **BMI (kg/m<sup>2</sup>)** | 28.6 ± 7.9 | 29.2 ± 7.3 | 0.26 |
| **Body Fat Percentage (%)<sup>A</sup>** | 31.3 ± 8.5 | 29.3 ± 7.5 | 0.13 |
| **VAT mass/total mass (%)<sup>A</sup>** | 0.6 ± 0.3 | 0.8 ± 0.3 | 0.03 |
| **Fasting glucose (mg/dL)** | 94 (87, 104) | 91.0 (86, 104) | 0.54 |
| **Fasting insulin (mU/L)<sup>A</sup>** | 10 (6, 17) | 10 (8, 17) | 0.66 |
| **HOMA-IR<sup>A</sup>** | 2.2 (1.3, 4.3) | 2.7 (1.6, 4.2) | 0.84 |
| **HbA1c (%)** | 5.6 ± 0.6 | 5.4 ± 0.5 | 0.11 |
| **Duration of HIV (years)** | - | 23 ± 11 | - |
| **Duration of ART use (years)** | - | 12 ± 7 | - |
| **CD4<sup>+</sup> cell count at enrollment (cells/mm<sup>3</sup>)** | - | 636 (515, 833) | - |
| **INSTI-based regimen, n (%)** | - | 39 (91%) | - |
| **Hydroxyproline (ng/mg fat)<sup>A</sup>** | 193.5 (59.6, 589.8) | 537.7 (301.7, 899.5) | <0.01 |

Values are presented as mean ± SD or median (interquartile range). Other category for race includes one individual identifying as American Indian, one as Pacific islander and three with unknown race. Differences between the groups were analyzed by Chi-Square test for categorical variables and Wilcoxon rank-sum test for continuous variables. <sup>A</sup> Missing data from n = 3 (Body fat percentage), n = 5 (VAT mass/total mass), n = 2 (Fasting Insulin and HOMA-IR) and n=6 (Hydroxyproline). PWH: people with HIV; PWoH: people without HIV; VAT: visceral adipose tissue; INSTI: Integrase Strand Transfer Inhibitor.

### PWH display excess SAT fibrosis, independent of overall body adiposity

We and others have shown that SAT fibrosis is closely linked to the development of IR, a major driver of T2D, in humans(24, 34, 35). Given their high risk for T2D, we therefore assessed levels of SAT fibrosis in PWH. To do so, we measured levels of hydroxyproline (HYP), a key component of collagen and a reliable marker of ECM remodeling(36), in SAT samples collected from our cohort. PWH had substantially higher HYP levels in the SAT than did PWoH (p<0.001, Figure 1A). Interestingly, whereas male sex (p < 0.001) and %BF (p < 0.001) were also significant predictors of SAT HYP levels, other demographic factors including race and age were not independently associated with SAT fibrosis in PWH. Indeed, the presence of HIV was an indicator of elevated SAT HYP content even after adjusting for sex, age, race/ethnicity and %BF, (p = 0.004), suggesting that ART-treated HIV infection is an independent driver of SAT fibrosis (Supplemental Table 2 and Supplemental Figure 1).

**Figure 1.**
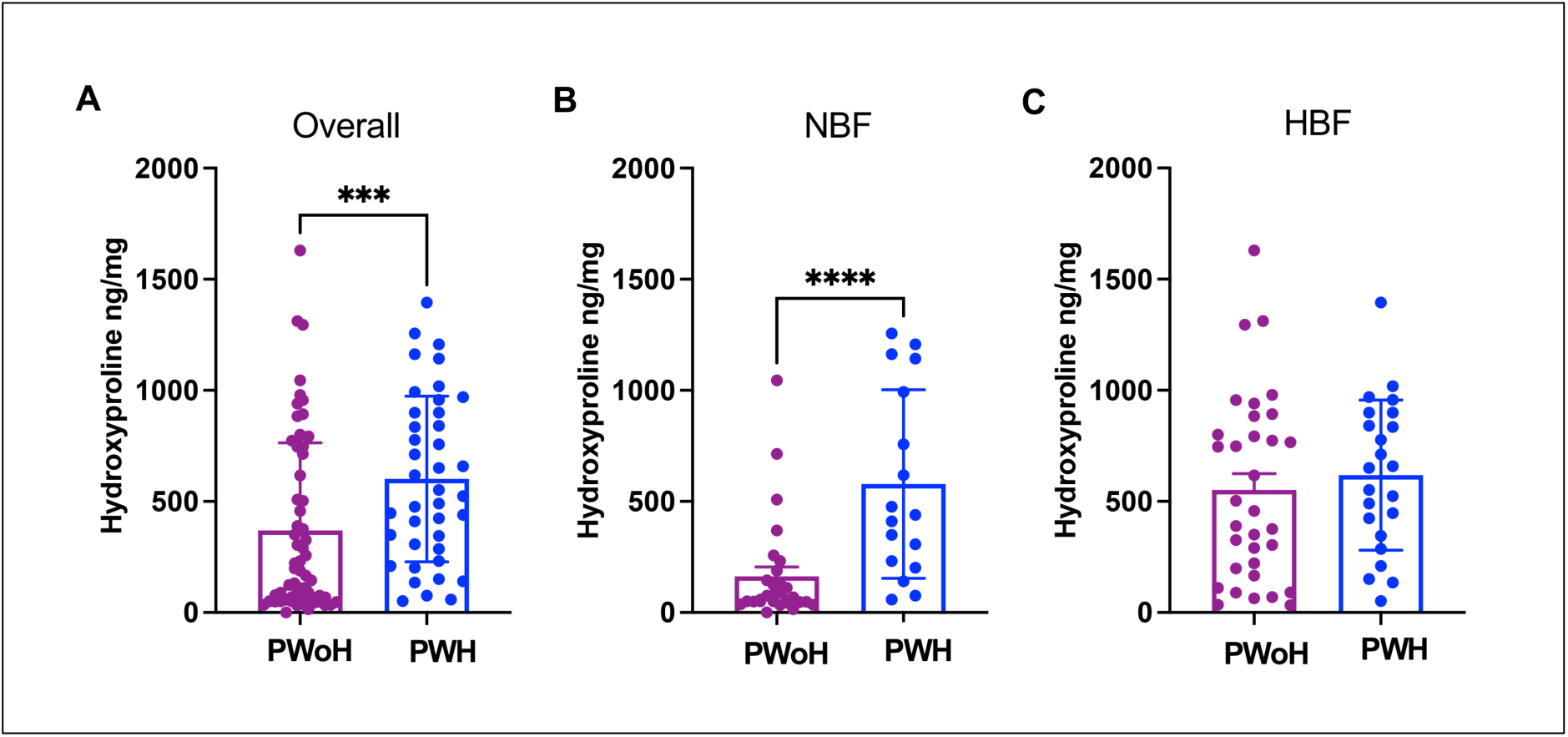
PWH have more AT fibrosis vs. PWoH, specifically in those with normal %BF. (A) SAT HYP levels from all study participants in the two groups (PWoH n = 62, PWH n = 40) showing higher HYP content per unit SAT in PWH (369.7 ± 393.7ng/mg for PWoH, 601.4 ± 372.5ng/mg for PWH, p = 0.0004). (B) Subgroup analysis for participants with normal %BF (PWoH n = 29, PWH n = 17) showing higher SAT HYP content in PWH with normal %BF. (162.5 ± 231.7ng/mg for PWoH, 578.1 ± 424.4 ng/mg for PWH, p < 0.0001) (C) Subgroup analysis for participants with high %BF (PWoH n = 33, PWH n = 23) showing no difference in SAT HYP content between the two groups (p = 0.34). Differences between the two groups were analyzed by Wilcoxon rank-sum test. *** p < 0.001, **** p< 0.0001

Given that increasing %BF and resultant obesity are both associated with the development of SAT fibrosis in the general population(24–26), we next performed a subgroup analysis after classifying the study participants into groups based on %BF. We chose to reanalyze by %BF, because it provides a more accurate assessment of whole-body adiposity than does BMI, which does not reliably differentiate between fat mass and lean mass(37), and because it has been shown to better mark metabolic disease risk vs. BMI(37–39). In this analysis, %BF values of ≥ 25% in males and ≥ 35% in females were used to define obesity, aligning with established cutoffs in the field(40–42). Accordingly, men were classified as having either “normal adiposity” (NBF, %BF < 25%) or “high adiposity” (HBF, %BF ≥ 25%), while females were classified as NBF if their %BF was <35% or as HBF if %BF was ≥ 35%. Remarkably, this subgrouping revealed that whereas levels of SAT fibrosis were not different between PWoH and PWH groups with HBF (Figure 1C), the relative increase in SAT fibrosis that we saw when analyzing PWH in aggregate was even more pronounced when the analysis was confined to those with NBF (p < 0.0001, Figure 1B). Together, these findings indicate that elevated SAT fibrosis in PWH is not driven by obesity but rather represents a distinct characteristic observable even in relatively lean individuals.

### SAT fibrosis is uniquely associated with markers of IR in nonobese PWH

To assess the potential implications of the marked difference in SAT fibrosis between PWH and PWoH on glycemic control, we assessed the correlation between SAT fibrosis, anthropometric measures, and HOMA-IR, a clinically useful marker of IR(43, 44), in both PWH and PwoH while adjusting for age, sex and race/ethnicity. In line with previous work from our group(24) and others(45, 46), increasing SAT fibrosis in PWoH correlated positively with increasing adiposity as assessed by both BMI and %BF and concomitantly with worsening IR, as measured by HOMA-IR (Figure 2A). In PWH, however, SAT fibrosis did not correlate with BMI or %BF, again supporting the concept that SAT fibrosis in this population is not simply a consequence of obesity (Figure 2B). Notably, while BMI and %BF were strongly linked to IR in PWoH, this relationship was absent in PWH, reinforcing the notion that factors beyond adiposity contribute to IR in this group. Despite the lack of correlation between SAT fibrosis and adiposity, SAT fibrosis in PWH remained positively associated with HOMA-IR, underscoring an obesity-independent contribution of SAT fibrosis to IR in this population.

**Figure 2.**
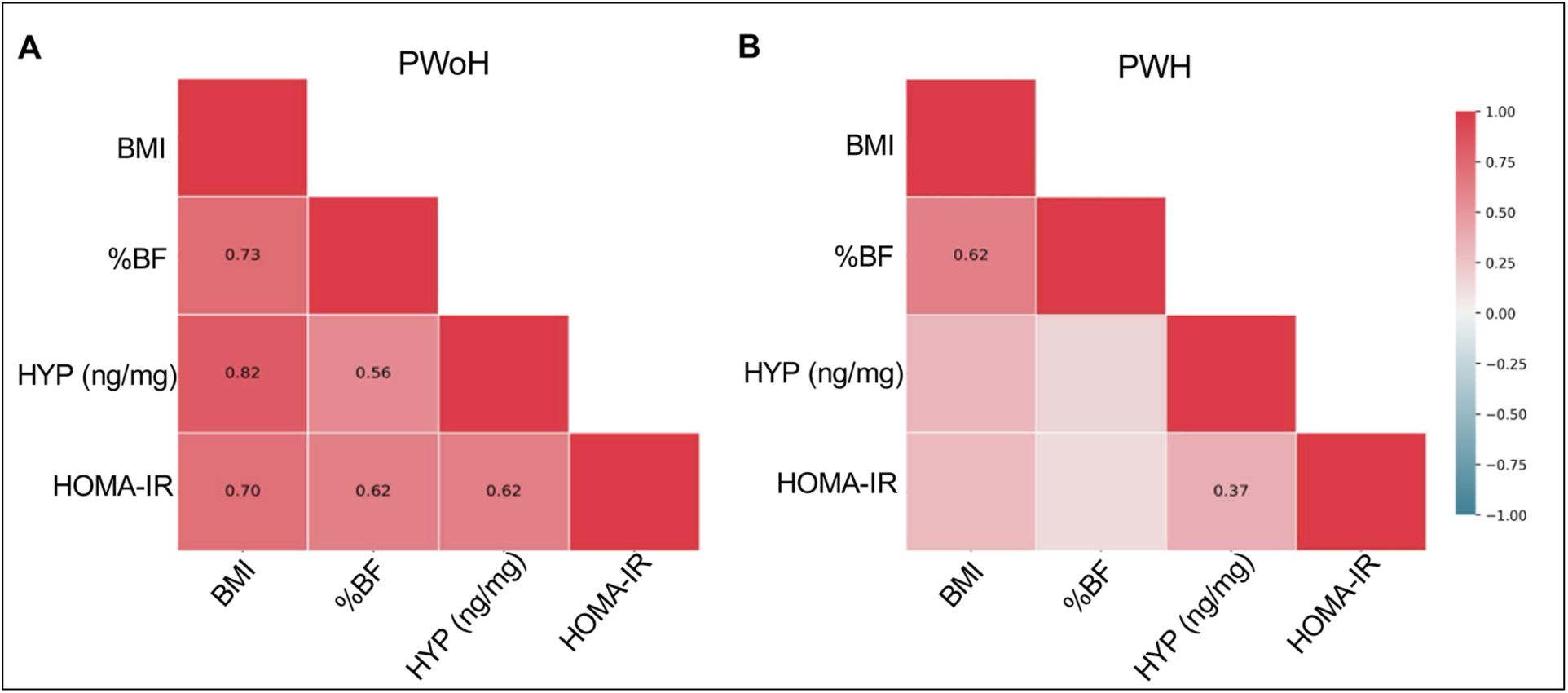
Relationships among BMI, total body fat percentage (%BF), hydroxyproline as a marker of SAT fibrosis, and insulin resistance measured by HOMA-IR. Panel A displays the correlation matrix for PWoH (n = 62), and Panel B shows the matrix for PWH (n = 40). Among participants without HIV, SAT fibrosis exhibited strong positive correlations with %BF, BMI, and insulin resistance. In PWH, the correlation between SAT fibrosis and insulin resistance was weaker, indicating distinct relationships between adiposity, SAT fibrosis, and insulin resistance in the two groups. Spearman’s rank-order correlation coefficients were calculated for each pair of variables while adjusting for the effects of sex, age, and race. The partial Spearman’s correlation coefficient rho is shown on the heatmap only for the correlations that meet statistical significance (p < 0.05). HYP: Hydroxyproline

To examine the impact of HIV status and adiposity on the relationship between SAT fibrosis and IR, participants were again categorized into four groups including PWoH-NBF, PWoH-HBF, PWH-NBF, and PWH-HBF using the same %BF cutoffs in male and female participants as previously defined. Among PWoH, SAT fibrosis correlated with HOMA-IR in the PWoH-HBF group (R = 0.46, p < 0.01) as expected, but not in the PWoH-NBF group (Supplemental Figure 2 A-B). PWH, by contrast, exhibited a reciprocal pattern, with SAT fibrosis strongly correlating with HOMA-IR in the PWH-NBF group (R = 0.70, p <0.05), but not in the PWH-HBF group. Together, these findings suggest that whereas the connection between SAT fibrosis and IR in PWoH is tightly associated with excess total body adiposity, SAT fibrosis in PWH contributes to the development of IR through mechanisms independent of adiposity, supporting the concept that SAT fibrosis specifically in relatively lean PWH could play a direct role in promoting IR.

### PWH have a distinct transcriptional pattern of fibrosis-associated gene expression in the SAT

Given the strong association between SAT fibrosis and IR in PWH even in the absence of obesity and despite the lack of correlation with increasing total body adiposity, we hypothesized that SAT fibrosis in PWH may be driven by distinct molecular pathways that are not engaged in the development of obesity-associated SAT fibrosis as seen in PWoH. To test this hypothesis, we performed transcriptomic profiling of SAT samples using a targeted panel of 772 fibrosis-related genes (Nanostring). Of the 112 participants in the cohort, 75 participants who provided sufficient SAT for mRNA isolation were included in the analysis. The characteristics of these participants are summarized in Supplemental Table 3.

Of the 772 fibrosis-related genes analyzed, a total of 45 genes were differentially expressed between PWH and PWoH (Figure 3). Of these, 34 genes were transcriptionally upregulated and 11 were downregulated, respectively, in PWH. Upregulated transcripts included those encoding for A) components of the ECM, such as collagen type 14 alpha 1 chain (*COL14A1*), fibril-associated collagen; elastin (*ELN*) which encodes a component of elastic fibers; and laminin subunit gamma 1 (*LAMC1*), which encodes the gamma-1 subunit of laminins (a family of ECM glycoproteins); and B) those encoding factors associated with ECM remodeling, such as inhibitors of matrix metalloproteinases [e.g. Tissue inhibitor matrix metalloproteinase inhibitor 1 (*TIMP1*) and 2 (*TIMP2*)]. The upregulation of *COL14A1* mRNA levels in the SAT of PWH is particularly noteworthy, as it reflects a discrepancy between the transcriptionally dominant collagen underlying SAT fibrosis in PWH (collagen type 14) vs. those found in the context of obesity-associated SAT fibrosis occurring among PWoH, in whom collagen type 14 is not well described and in whom collagens type 1, 3, 4 and 6 transcriptionally predominate(27, 34, 45).

**Figure 3.**
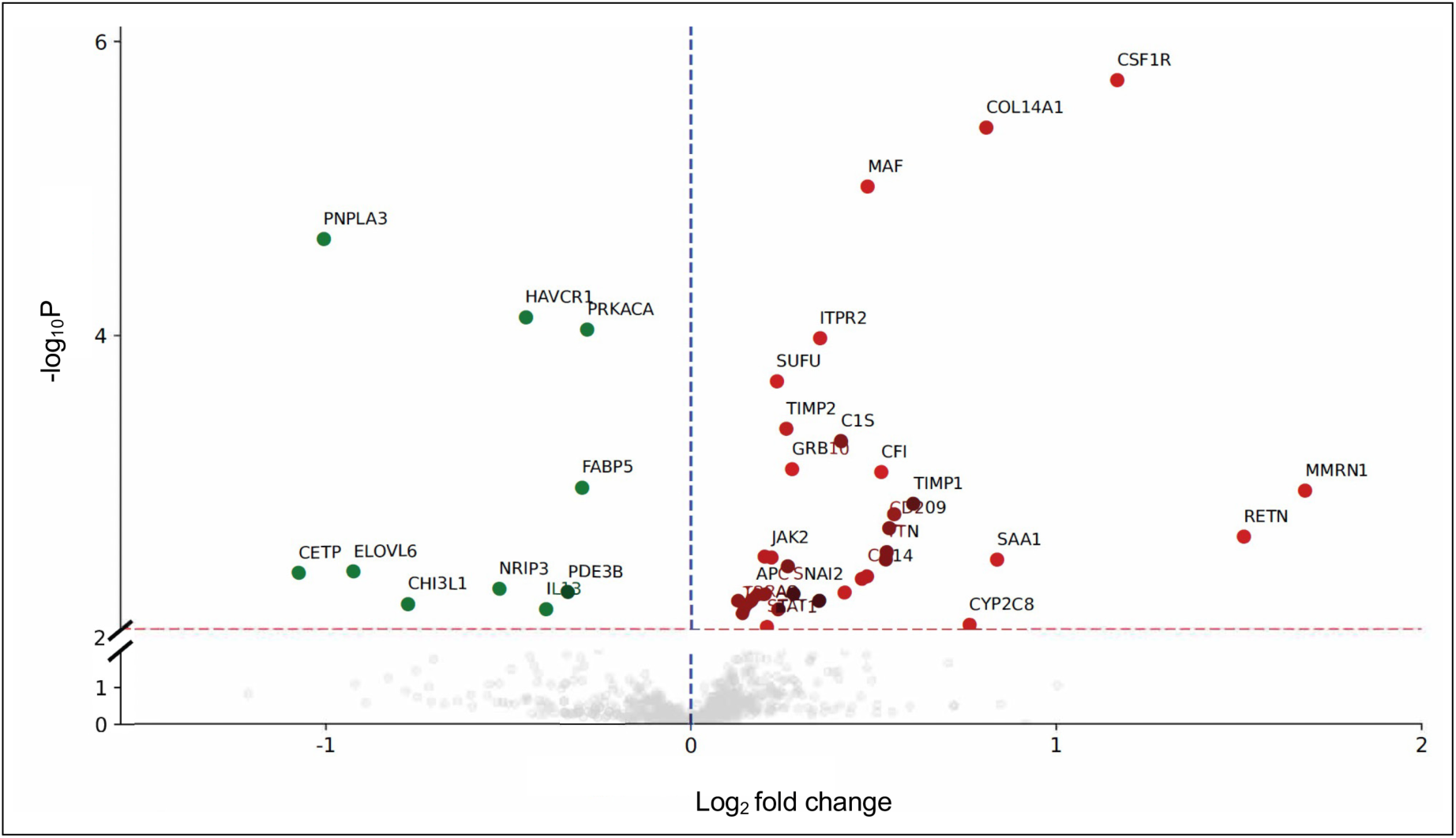
PWH Have a Distinct Transcriptional Pattern of Fibrosis-associated Gene Expression in The SAT. Volcano plot showing differentially expressed genes in the SAT of PWH (n = 37) vs. PWoH (n = 38). Dashed lines indicate log_2_ fold change (blue) and a p-value cutoff of 0.01 (red). Significantly upregulated and downregulated genes are represented by green and red dots, respectively. Differential expression was measured using generalized linear models of the negative binomial family, adjusted for age, sex, race, ethnicity, %BF, and batch.

Gratifyingly, several genes involved in immune signaling were transcriptionally upregulated in the SAT of PWH, both validating our approach and further highlighting the impact of HIV infection on AT. These included genes encoding proteins critical for pathogen recognition and antigen presentation, such as *CD14*, a co-receptor for bacterial lipopolysaccharide (LPS) expressed on myeloid cells that drives immune activation and inflammation(47), and *CD209*, a C-type lectin receptor essential for pathogen binding, with polymorphisms associated with susceptibility to HIV-1 infection(48, 49). These findings reinforce the concept that HIV infection induces widespread immunologic alterations beyond traditional immune compartments, extending to AT and aligning with emerging evidence from other studies.

By contrast, genes that were transcriptionally downregulated in PWH compared to PWoH include those encoding proteins involved in lipid handling, such as cholesteryl ester transfer protein (*CETP*), fatty acid elongase 6 (*ELOVL6*), fatty acid binding protein 5 (*FABP5*), and patatin-like phospholipase domain-containing 3 (*PNPLA3*), which encodes a triacylglycerol (TG) lipase responsible for TG hydrolysis. This pattern of transcriptional downregulation suggests that the pro-fibrotic and pro-inflammatory shifts in the SAT of PWH occur at the expense of its lipid metabolic functionality, which is critical for normal AT health.

### The HIV-specific impact on SAT gene transcription is distinct from that of obesity

Since HIV and obesity had distinct effects on SAT HYP levels, we next sought to determine whether they also had distinct effects on SAT transcriptional profiles. We therefore stratified our transcriptional dataset to delineate the transcriptional impact of HIV infection on fibrosis-related gene expression in SAT from the effects of increasing adiposity independent of HIV. Study participants were once again categorized into four groups—PWoH-NBF, PWoH-HBF, PWH-NBF, and PWH-HBF—mirroring the stratification used in earlier analyses of the relationship between SAT fibrosis and HOMA-IR. The PWoH-NBF group served as the reference group for differential gene expression (DGE) analyses (Figure 4A and Supplemental Figure 3), the results of which we summarized as three comparative pairwise analyses in a Venn diagram (Figure 4B and Supplemental Table 4). The number of genes downregulated in PWoH-HBF, PWH-NBF, and PWH-HBF, respectively, when compared with PWoH-NBF participants in a pairwise manner is shown, with an analogous approach used to depict the upregulated genes for each of the same pairwise comparisons (Figure 4B).

**Figure 4.**
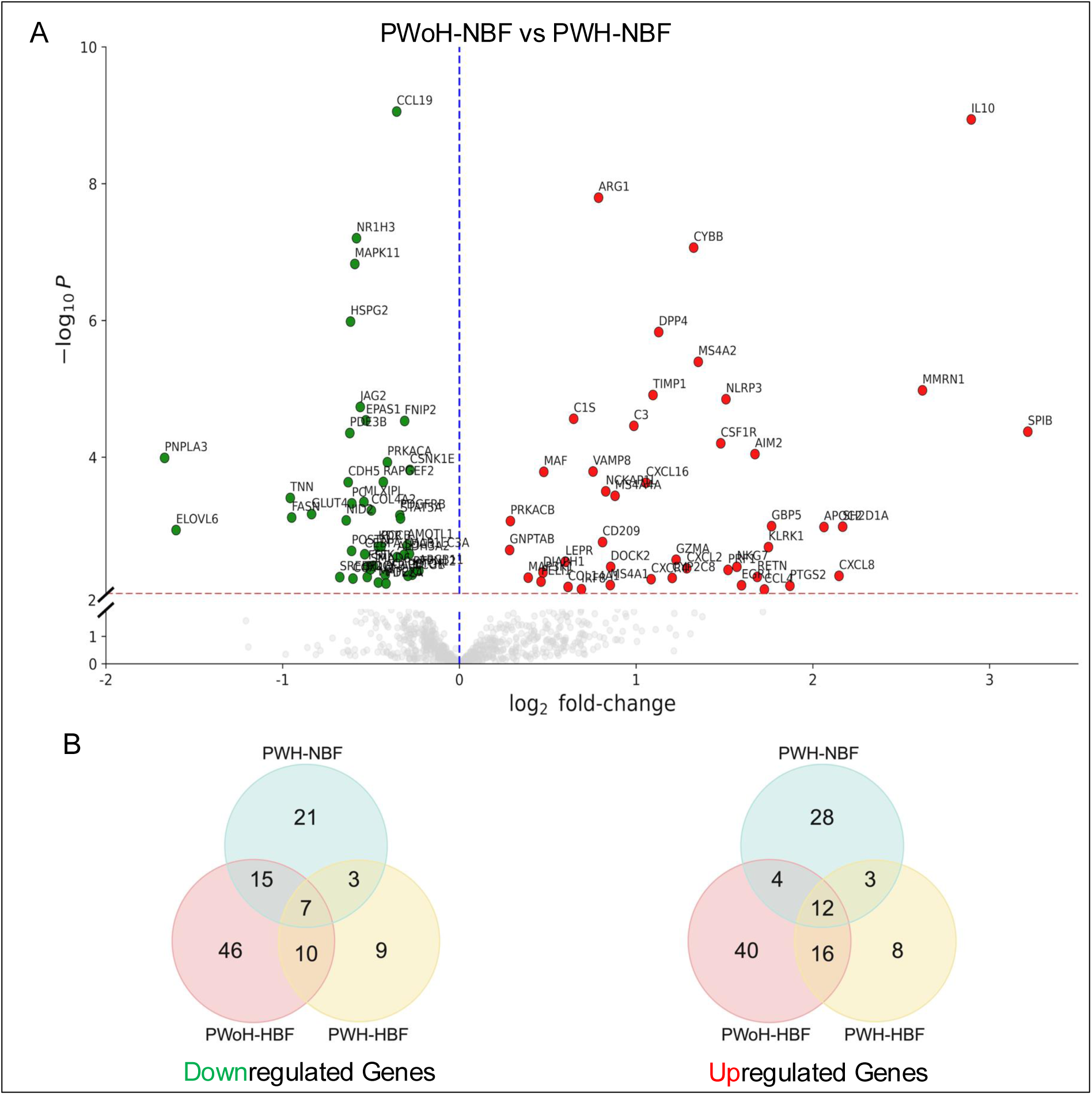
The HIV-Specific Impact on SAT Gene Transcription is Distinct From that of Obesity. (A) Volcano plot showing differentially expressed genes (DEGs) in the SAT from PWH-NBF (n = 16) compared to the control group, PWoH-NBF (n = 18). DEGs were identified using linear modeling (limma), with a nominal p-value cutoff of 0.01 and log₂ fold-change thresholds. Genes with increased expression in each comparison are shown in red and those with decreased expression in green. (B) Venn diagrams summarizing the total number of upregulated and downregulated genes in each of the three pairwise comparisons vs. PWoH-NBF. Overlapping regions indicate genes shared across multiple comparisons, highlighting distinct and convergent transcriptional responses to HIV infection and increased adiposity.

Reassuringly, our DGE analysis recapitulated many transcriptional changes known to occur in the SAT of people responding to increased adiposity in the general population^51^. For example, the comparison of PWoH-HBF vs. PWoH-NBF (control) groups revealed several upregulated genes already reported to be induced by obesity, including those encoding inflammatory markers (*CCL13*, *CCL19*) and factors involved in ECM remodeling (LOX, LOXL1, LOXL4, TIMP2)(50, 51) (Supplemental Figure 3, Table 2). Analysis of downregulated genes similarly confirmed well established findings in the literature(51–53), as mRNA levels of genes encoding adiponectin (*ADIPOQ*), the insulin receptor (*INSR*), insulin receptor substrate 1(*IRS1*), and phosphatidylinositol-4,5-bisphosphate 3-kinase catalytic subunit alpha (*PIK3CA*), were all downregulated in PWoH-HBF vs. PWoH-NBF. Together, these confirmatory findings provide us with confidence in our cohort, tissue collection and processing methodology, and analytical pipeline.

**Table 2.**
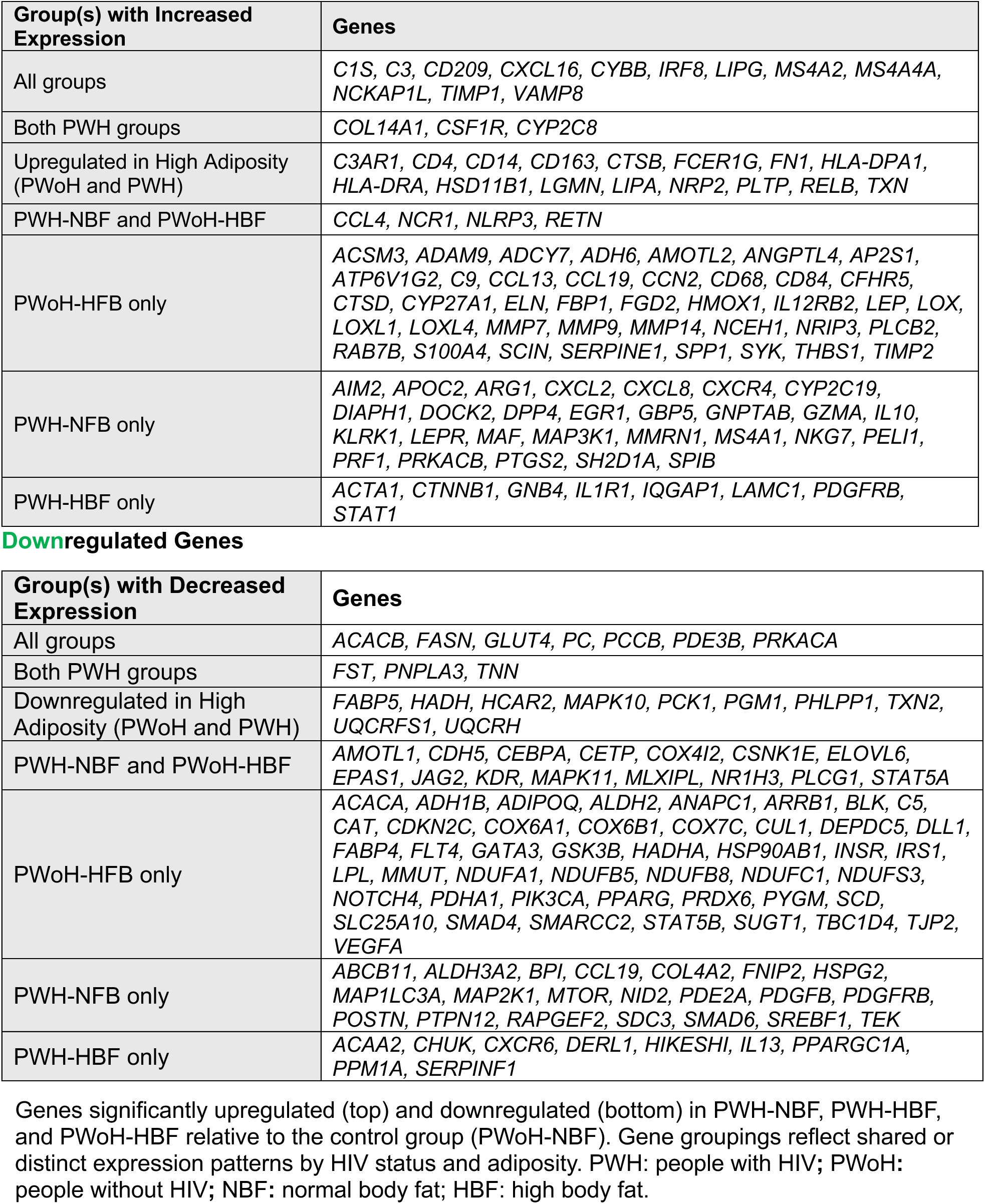
Differentially expressed genes in adipose tissue across study groups stratified by HIV status and body fat. **Upregulated Genes**

### Identifying a fibrosis-related transcriptional signature unique to HIV infection and independent of adiposity

From these three sets of pairwise DGE analyses, we next focused on genes that were differentially regulated at the intersection of two or all three of the subgroups. Analyzing the DGEs found at these intersections (Figures 4B and Table 2) revealed mRNAs upregulated across each of the three DGE analyses we performed vs. the reference PWoH-NBF group. Three genes (*COL14A1*, *CSF1R*, and *CYP2C8*) were upregulated in both the PWH-NBF and the PWH-HBF groups vs. the PWoH-NBF reference group, indicative of genes that are transcriptionally induced in the SAT of PWH regardless of total body adiposity. By contrast, four other genes (*CCL4, NCR1, NLRP3*, and *RETN*) were transcriptionally upregulated in both the PWH-NBF and PWoH-HBF subgroups vs. the reference group, indicative of genes involved both in the response of PWoH to increasing total body adiposity and also in the response of relatively leaner participants to HIV infection. This overlapping set of genes highlights the possibility that some components of the SAT fibrosis seen in PWH-NBF results from an engagement of responses that are also induced by obesity in the general population. Furthermore, sixteen genes were transcriptionally upregulated in the SAT of both PWoH-HBF and PWH-HBF participants compared to the reference group (PWoH-NBF), highlighting genes that are induced by increasing total body adiposity, regardless of HIV infection status.

Our parallel analysis of downregulated genes was also revealing (Figure 4B, and Table 2). Specifically, 7 genes were transcriptionally downregulated in all three of the pairwise comparisons made vs. the PWoH-NBF group. These notably include solute carrier family 2 member 4 (*GLUT4*), which encodes the glucose transporter responsible for insulin-mediated glucose uptake in adipose tissues, a function that is impaired in the context of IR. 3 genes (*FST*, *PNPLA3*, and *TNN*) were transcriptionally downregulated in both PWH-NBF and PWH-HBF participants vs. the PWoH-NBF reference group, reflecting transcriptional responses to HIV infection regardless of total body adiposity. Another 10 genes were downregulated in both PWH-HBF and PWoH-HBF groups, highlighting pathways repressed by increasing total body adiposity independently of the presence or absence of chronic HIV infection. These genes include those encoding proteins associated with intracellular lipid handling (*FABP5*), fatty acid β-oxidation (hydroxyacyl-CoA dehydrogenase *HADH*), mitochondrial membrane potential regulation (thioredoxin 2 *TXN2*), and mitochondrial respiratory electron transport (ubiquinol-cytochrome c reductase Rieske iron-sulfur polypeptide 1 *UQCRFS1*, ubiquinol-cytochrome c reductase hinge protein *UQCRH*) that are critical for the normal metabolic functions of ATs. In all, our granular subgroup analyses revealed both the mechanistically separate and shared transcriptional impacts of both chronic, treated HIV infection and obesity on the transcription of genes across a large panel of fibrosis-related genes in the SAT.

Functional pathways corresponding to genes differentially expressed in the SAT were identified using Gene Set Enrichment Analysis (GSEA) with curated databases including the Kyoto Encyclopedia of Genes and Genomes (KEGG), Reactome, and Gene Ontology (GO) gene sets. KEGG and Reactome provide pathway-based maps describing molecular interactions and biological processes, while GO offers gene classifications based on molecular function, biological processes, and cellular components. This analysis enabled us to delineate biological processes that might distinguish the excess SAT fibrosis in PWH-NBF from those that are operational in both PWH-HBF and PWoH-NBF, highlighting potential pro-fibrotic transcriptional mechanisms in PWH that manifest independently of obesity.

In PWH-NBF, pathways related to ECM remodeling, immune activation, and leukocyte migration were significantly upregulated, consistent with a pro-fibrotic and inflammatory SAT phenotype (Figure 5A). Concurrently, there was significant downregulation of pathways related to insulin signaling, thermogenesis, and lipid metabolic processes. Together, these results suggest that SAT dysfunction in PWH-NBF is characterized by immune- and matrix-driven remodeling and impaired energy metabolism, even in the absence of obesity. In PWH-HBF, ECM and immune-related pathways remained upregulated, overlapping with those observed in PWH-NBF, but with additional enrichment of cytokine signaling, complement activation, and antigen presentation pathways, suggesting that adiposity may further amplify inflammation and immune dysregulation in the setting of HIV (Figure 5B). As expected, and previously shown in obesity-related studies, PWoH-HBF SAT also demonstrated upregulation of ECM pathways and downregulation of thermogenesis and adipocytokine signaling, consistent with established features of AT dysfunction in obesity alone (Figure 5C). These findings support the concept that while obesity independently contributes to fibrotic and metabolic remodeling, ART-treated HIV introduces additional immune perturbations that distinguish the SAT environment in PWH from PWoH, underscoring the potential need for alternative metabolic risk assessment and therapeutic strategies that specifically target ECM remodeling and immune dysregulation in PWH.

**Figure 5.**
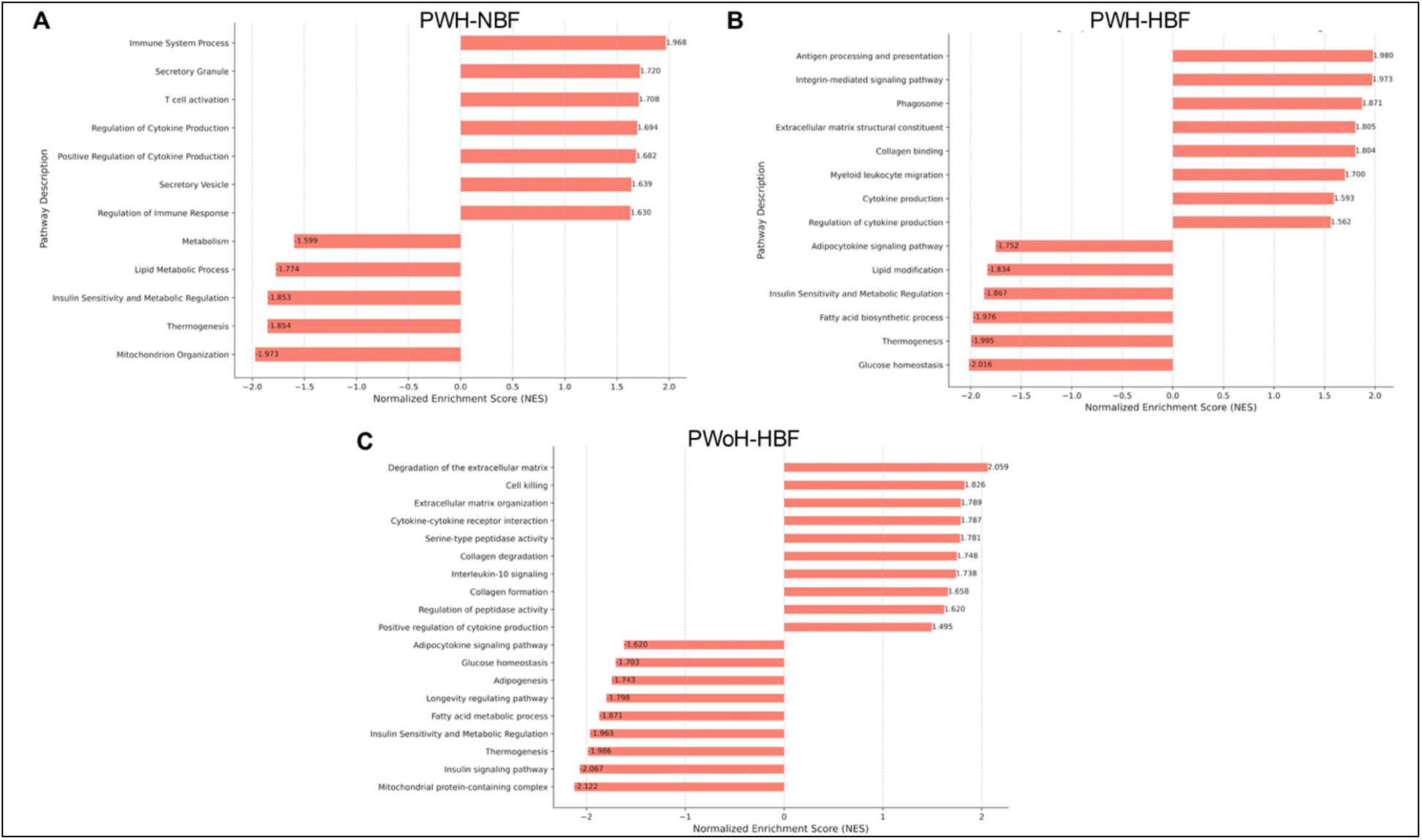
Pathway analysis reveals downregulation of insulin sensitivity, thermogenesis, and lipid metabolic pathways in SAT from PWH with normal adiposity, alongside shared enrichment of inflammatory and ECM remodeling programs across groups. Normalized enrichment score plots of pathways significantly enriched in PWH-NBF (A), PWH-HBF (B) and PWoH-HBF (C), versus PWoH-NBF (control group) using GSEA analysis (false discovery rate cutoff for pathway = 0.1).

### Endotrophin is an adiposity-independent mediator of SAT Fibrosis in PWH

We next sought to determine if SAT fibrosis in PWH can be marked in a manner that does not require an invasive tissue biopsy. To this end, we focused on endotrophin (ETP), a cleavage product of collagen 6A3 that promotes ECM remodeling, immune cell infiltration, and fibrotic signaling in the SAT, contributing to increased tissue stiffness and metabolic dysfunction(26, 54, 55). Moreover, there is interest in targeting ETP and downstream signaling as a means to reestablish metabolic health and glycemic control in the context of metabolically unhealthy obesity(55, 56). However, ETP levels have not been assessed in PWH to our knowledge.

As a foundational step, we examined SAT gene expression from our NanoString dataset, comparing PWH to PWoH. We assessed expression of *COL1A1*, *COL1A2*, *COL3A1*, and *COL6A3*, all of which have been previously associated with AT fibrosis in human obesity. Although differences between groups were not statistically significant, we observed a consistent trend toward higher expression of all four genes in PWH (Supplemental Figure 4). These findings suggest that beyond collagen 14a1, increased fibrotic remodeling in the SAT of PWH may involve multiple collagens and offers support for examining ETP as a potential circulating marker of tissue fibrosis in this population.

We found that, as expected, PWoH-HBF had higher plasma ETP levels than did individuals in the PWoH-NBF group, consistent with prior reports linking ETP to obesity and metabolic dysfunction in the general population (Supplemental Figure 5). Remarkably, however, we also found that plasma ETP levels were robustly elevated in PWH vs. PWoH, even after adjusting for sex, age, and race/ethnicity (Supplemental Figure 5A). Moreover, when participants were stratified by %BF into NBF and HBF groups, ETP levels remained significantly higher in PWH-NBF vs. PWoH-NBF individuals (p <0.01), supporting the notion that factors beyond adiposity contribute to ETP elevation in relatively lean PWH and mirroring our findings when measuring HYP levels in the SAT itself (Supplemental Figure 5B). Indeed, ETP levels multivariable regression models were a significant independent predictor of SAT fibrosis in PWH (p = 0.013, Supplemental Table 5), even after adjusting for age, sex, race/ethnicity, and % body fat. These findings suggest that HIV infection is independently associated with elevated circulating ETP levels, particularly among those in the NBF group, and that plasma ETP levels may serve as a noninvasive biomarker of SAT remodeling among individuals in this population, with potential implications for metabolic risk stratification and therapeutic targeting.

## Discussion

In this study, we demonstrate that PWH exhibit increased SAT fibrosis compared to those without HIV, occurring in a manner uncoupled from obesity. In PWH-NBF, SAT fibrosis showed a strong association with IR, whereas commonly used anthropometric measures such as BMI and %BF were not similarly predictive. These findings position SAT fibrosis as a critical driver of metabolic dysfunction in PWH, independent of obesity, and emphasize the need for alternative metrics to assess metabolic risk in this population.

Current strategies to mitigate the increased risk of T2D in PWH focus primarily on routine screening and lifestyle interventions, similar to those applied in PWoH. The American Diabetes Association guidelines recommend screening prior to initiating ART, followed by annual follow-ups to facilitate early diagnosis. However, the underlying mechanisms contributing to this heightened risk remain unclear. Our data highlight the limitations of relying on BMI as a metric for assessing metabolic risk in PWH, particularly in non-obese individuals, and underscore instead the importance of understanding the unique metabolic pathways in PWH to develop tailored risk assessment tools and potentially even targeted therapies.

Few studies have previously explored the role of SAT fibrosis in metabolic health among PWH. Bailin et al.(57) reported transcriptional and cellular changes in the SAT from a cohort of PWH with varying T2D status—including individuals without diabetes, with prediabetes, and with T2D implicating pathologic fibroblasts in the accrual of VAT and the development of T2D. However, the lack of a control group of PWoH in their study limited the ability to identify HIV-specific effects. In contrast, our study provides direct comparisons between PWH and PWoH, including stratification by adiposity, which enabled us to discern HIV-specific transcriptional changes from those driven by obesity per se. Importantly, our measurements of HYP content in the SAT confirm that transcriptional upregulation of ECM genes translates into actual protein-level changes, strengthening the link between SAT fibrosis and IR in PWH.

Our findings align with prior research showing the association between SAT fibrosis and metabolic dysfunction in the general population(24, 27, 34). Collagens type 1, 3, 4, and 6 dominate in pathological adipose tissue and contribute to AT dysfunction(27, 34, 45). However, whereas there was a trend towards upregulation of these collagen genes in PWH, we identified COL14A1 as being uniquely upregulated in PWH, independent of excess adiposity. Collagen type 14, a fibril-associated collagen with interrupted triple helices (FACIT), which consists of two collagenous domains, which are important for the formation of the COL14 triple helical structures and for the binding to other fibrillar collagens, and three non-collagenous domains(58, 59). COL14 has been implicated in cell proliferation, differentiation and fibrillogenesis and it has been identified in various tissues, including skin, tendon, cornea, and lung, where it localizes near blood vessels, airway smooth muscle, and bronchial epithelium(60–62). While collagen type 14 function in ECM remodeling and mechanical stress regulation is well-established in these tissues, very few studies have reported on *COL14A1* in AT and its contribution to AT structure, fibrosis, and metabolic dysfunction remains largely uncharacterized. In our study, COL14A1 was upregulated in PWH compared to PWoH, suggesting a that it may have a distinct role in SAT remodeling in the context of HIV. A microarray analysis previously found that *COL14A1* expression decreases during adipogenic differentiation of human mesenchymal stem cells(63), whereas in 3T3-L1 cells, collagen 14—particularly its FN type III domain—was shown to promote adipogenic differentiation(64). A recent study found that *COL14A1* mRNA expression was elevated in PWH with increased VAT volume(57), an interesting corroboration of our findings. Together, these data suggest that *COL14A1* may contribute to fibrotic remodeling in adipose tissue, particularly in PWH, where SAT fibrosis is associated with insulin resistance and metabolic complications.

Also transcriptionally upregulated in the SAT of PWH vs. PWoH were genes related to cellular processes known to promote tissue fibrosis in general. For example, *TIMP1* levels were also shown to be induced in in vitro models of human metabolic-dysfunction associated steatohepatitis (MASH)-associated liver fibrosis(65). Our analysis also showed upregulation of the gene encoding Janus kinase 2 (JAK2), which was reported to mediate IL-5a-dependent pro-fibrotic signaling from in the setting of lung fibrosis(66). Importantly, JAK2 signaling is also important in response to inflammatory cytokines known to be elevated in HIV including interferon-γ and IL-6. Collectively, these findings suggest that chronic HIV infection transcriptionally induces pathways in the SAT that mediate fibrosis in multiple tissue environments.

Additionally, our transcriptional analysis revealed overlapping pathways between HIV infection and obesity. For example, *CCL4* and *NLRP3* were upregulated in both PWH and participants with high adiposity. *CCL4* plays a dual role in HIV suppression and metabolic inflammation, while *NLRP3* is a key component of the inflammasome implicated in adiposity and HIV-related inflammation(67–69). These overlapping transcriptional responses suggest shared mechanisms contributing to insulin resistance and T2D risk in PWH, warranting further exploration of their cellular origins and functional relevance.

In terms of clinical implications, the pathways identified in this study—particularly those related to impaired adipogenesis and excessive ECM remodeling—underscore the need to move beyond weight loss–focused strategies and instead consider interventions that target adipose tissue structure and function. Our findings support the concept that SAT fibrosis contributes to metabolic risk independent of adiposity, especially in PWH. This points to a potential role for therapeutic strategies aimed at improving adipose tissue expandability and reducing fibrotic remodeling. Anti-fibrotic agents currently in development for conditions such as liver fibrosis and NASH, including inhibitors of TGF signaling or LOXL2, may eventually have relevance in the context of adipose tissue fibrosis. Further research is needed to evaluate whether such therapies could benefit metabolic health in PWH by mitigating fibrosis-driven insulin resistance.

Building on these findings, we focused on endotrophin (ETP), a circulating cleavage product of *COL6A3*, known to be associated with tissue remodeling, fibrosis, and metabolic dysfunction. ETP has been shown to promote inflammation and insulin resistance in adipose tissues, where its levels increase with obesity and associated adipose tissue fibrosis(26). Human studies have also revealed a significant upregulation of ETP in the adipose tissues of individuals with obesity and T2D(70, 71). Indeed, ETP is also increasingly being recognized as a key mediator of inflammation and fibrosis in the pathophysiological remodeling seen in both metabolic and other pro-fibrotic diseases(72). ETP is highly expressed in tumors and accelerates cancer progression, including breast and liver cancer(73, 74). Clinically, elevated circulating ETP is associated with poor outcomes in chronic kidney disease(75), chronic liver disease(76) and has been identified as a predictive marker for response to insulin-sensitizing therapies(70). Our findings here go beyond obesity-associated fibrosis to suggest a distinct contribution of ETP to adipose tissue remodeling in PWH, independent of increased adiposity. The elevation of plasma ETP in PWH, particularly in those with NBF, supports the idea that HIV-related factors, rather than obesity alone, drive fibrosis in this population. Given that ETP is a circulating marker, it holds potential as a non-invasive biomarker for SAT fibrosis and its associated metabolic consequences in PWH. Future studies should explore whether targeting ETP-mediated fibrosis could mitigate metabolic dysfunction in HIV, providing a potential therapeutic avenue for this high-risk group.

Despite its strengths, our study also has limitations. While we aimed to balance age and sex between groups, practical recruitment challenges made complete demographic matching difficult. Specifically, it was challenging to recruit women with HIV despite targeted outreach, resulting in a PWH group that included a higher proportion of males and a greater proportion of Black participants—reflecting the real-world demographics of populations most at risk for HIV infection and its metabolic complications. Although we adjusted for age, sex, race/ethnicity, and percent body fat in our analyses, some residual confounding may remain, as is common in cross-sectional studies. However, in linear regression models, HIV status remained a significant independent predictor of SAT fibrosis even after adjusting for these variables, supporting the robustness of this association. Our transcriptional analysis was limited to a fibrosis-focused gene panel, potentially excluding other relevant pathways. Moreover, because we assessed bulk transcriptomics, we cannot differentiate altered gene expression within a given cell type from a difference in the admixture of cells with different transcriptional profiles. For example, reduced lipid metabolism gene expression in PWH may reflect an increased relative abundance of immune cells in the adipose tissue and a reduced abundance of adipocytes rather than a direct effect on adipocyte function. Furthermore, as the majority (91%) of the PWH in our study were on integrase strand transfer inhibitor (INSTI)-based regimens, it is not possible to disentangle the metabolic and transcriptional effects of HIV infection itself from potential direct effects of INSTI therapy. Finally, the cross-sectional nature of this study also precludes causal inferences. Longitudinal studies will be essential to clarify temporal relationships and to determine whether SAT fibrosis and plasma ETP levels predict long-term metabolic outcomes in PWH. Future research should also prioritize the recruitment of underrepresented groups—particularly women with HIV—to improve demographic matching and reduce potential confounding in comparative analyses.

In conclusion, our findings reveal increased SAT fibrosis in PWH, particularly in non-obese participants, and demonstrate its strong association with insulin resistance, independent of obesity. The distinct transcriptional profile of SAT fibrosis in PWH, including the upregulation of COL14A1, highlights unique HIV-driven mechanisms contributing to adipose tissue dysfunction. The elevation of plasma ETP levels in PWH further supports its potential as a biomarker and therapeutic target. These findings challenge the reliance on traditional anthropometric measures for assessing metabolic risk in PWH and provide a foundation for future studies to elucidate the cellular and molecular drivers of HIV-associated SAT fibrosis and its metabolic consequences.

## Methods

### Sex as a biological variable

Our study examined male and female participants. Sex differences are reported and were considered as a biological variable.

### Participants and enrollment procedures

We enrolled people with and without HIV aged 18–75 years from two UCSF-based cohorts: the SCOPE cohort and the Inflammation, Diabetes, Ethnicity, and Obesity (IDEO) cohort. The SCOPE cohort is a comprehensive longitudinal study of PWH and PWoH, focusing on detailed clinical and virologic outcomes. For this study, we included SCOPE participants who had been on ART for at least one year and maintained viral suppression with undetectable plasma HIV RNA levels for at least 12 months. The IDEO cohort comprises people without HIV recruited from UCSF and Zuckerberg San Francisco General Hospital medical and surgical clinics, as well as through local public advertisements. Established members of the IDEO cohort who met study criteria were included. Exclusion criteria for both cohorts included a prior diagnosis of T2D (HbA1c >6.5% or a physician diagnosis with diabetes medication use), use of anti-inflammatory medications or glucocorticoids, a history of organ failure, autoimmune disorders, cancer, or significant weight change (>5%) in the preceding three months. We stratified our sampling by HbA1c to ensure a comparable distribution of participants with prediabetes in each group, allowing us to assess whether the associations between adipose tissue and insulin resistance differed in the context of HIV status.

### Body composition and anthropometric measurements

Participants’ height and weight were measured using a standard stadiometer and scale, with body mass index (BMI) (kg/m2) calculated from two averaged measurements. Body composition was estimated by dual-energy X-ray absorptiometry (DXA) using a Hologic Horizon/A scanner (3-minute whole-body scan, <0.1 G mGy) per manufacturer protocol. This device accurately measures participants up to 450 lbs and employs high-performance and “offset” scanning techniques on those whose bodies are wider than the table. Standard DXA readouts include total percentage body fat (%BF), fat mass, lean mass, total body mass and VAT volume. A single technologist analyzed all DXA measurements using Hologic Apex software (13.6.0.4:3) following International Society for Clinical Densitometry guidelines, with precision error (1 SD) for total fat mass and %BF of approximately 0.3 kg and 1%, respectively (calibration using device-specific whole-body phantoms). VAT was measured in a 5-cm-wide region across the abdomen just above the iliac crest, coincident with the fourth lumbar vertebrae, to avoid interference from iliac crest bone pixels and matching the region commonly used to analyze VAT mass by computed tomography (CT) scanning(77). Determination of VAT mass by DXA has been validated vs. CT as a reference standard in subjects with BMI between 18.5 and 40 kg/m2, with DXA and CT data correlating with a coefficient of determination (r2) of 0.959 (female) and 0.949 (men)(78).

### Adipose tissue sample collection

Subcutaneous adipose tissue (SAT) samples were collected from most participants after a 10-hour fast via aspiration biopsy under local anesthesia using 2% lidocaine. A 2.1 mm blunt-sided, ported liposuction catheter (Tulip CellFriendly™ GEMS Miller Harvester, Tulip Medical Products, San Diego, CA) was used to collect tissue from approximately 5 cm lateral to the umbilicus. In some participants, SAT was instead collected intraoperatively during bariatric surgery. After removal of visible connective tissue, blood, and clots, samples were washed with Krebs-Ringer bicarbonate buffer containing 1% BSA and immediately frozen in liquid nitrogen.

### Collagen content measurement

The collagen content in SAT was measured by assessing the levels of hydroxyproline (Hyp) determined by the Quick Zyme Sensitive Tissue Hydroxyproline Assay (QuickZyme Biosciences, the Netherlands) according to the manufacturer’s instructions. Briefly, the adipose tissue was acid hydrolyzed by 6M HCI at 95°C for 20 hours. The hydroxyproline content was then assessed by spectrophotometry at 570nm.

### ELISA based measurements of circulating endotrophin

Plasma samples were freshly prepared from blood samples collected after a 10 hour fast and preserved at –80°C until analysis. To quantitatively determine the levels of circulating endotrophin, 96-well plates (Corning Costar) were coated with rabbit monoclonal anti-endotrophin antibodies prepared in house at 2 μg/mL concentration. Plasma samples were titrated at a series of dilutions in 1X PBS, then added to an anti-endotrophin coated plate. A high affinity specific anti-endotrophin antibody (ETN-1Rb) was utilized as secondary detection antibody. Anti-Rabbit Fab2-HRP antibody (Jackson ImmunoResearch) was used for detection of endotrophin signals, using the dilution as suggested by the manufacturer. A purified endotrophin recombinant protein was titrated in a series of concentrations (0–50ng/mL) to establish a standard curve for calculation of endotrophin in plasma samples.

### RNA isolation and transcriptional analysis

Total RNA was extracted from 70 to 100 mg of whole SAT using the RNeasy Lipid Mini Kit (QIAGEN, Valencia, CA) following the manufacturer’s protocol. RNA concentration (ng/μL) and purity (A260/280) were determined for each RNA sample using the Nanodrop 1000 (ThermoFisher Scientific, Wilmington, DE). Individual mRNA levels and the transcriptional patterns for an array of fibrosis-associated genes in the SAT were analyzed using the NanoString customized fibrosis transcriptomic panel, consisting of 14 probes for quality control (6 positive controls, 8 negative controls), 8 probes for predetermined housekeeping genes (*ACAD9, ARMH3, CNOT10, GUSB, MTMR14, NOL7, NUBP1*, and *HPRT1*), and 772 probes for endogenous genes known to be involved in tissue fibrosis and the ECM. Raw counts were first filtered through a quality control step utilizing the 14 synthetic probes, then normalized to the mRNA levels of the housekeeping genes. The R package NanostringDiff was used for differential gene expression (DGE) analysis of the normalized counts. DGE analysis between PWH and PWoH was adjusted for age, sex, race, ethnicity, %BF and batch.

### Statistics

Statistical analyses were carried out in Python except for those pertaining to SAT gene expression analysis as described above. To compare the basic characteristics of PWH with PWoH, chi-square test was used for categorical variables and Wilcoxon rank-sum test was used for continuous variables. When analyzing transcriptional data, we followed the recommendations provided by the Nanostring manufacturers and did not implement traditional p-value adjustment approaches to account for multiple hypothesis testing in the DGE analysis. The reason for this is that the target genes in the fibrosis panel are not randomly selected. Instead, we applied a stringent p-value cutoff (p < 0.01) to identify genes with significant differential expression. To distinguish the transcriptional patterns associated with HIV infection from those associated with adiposity, study participants were categorized into four groups based on HIV status and %BF. A %BF cutoff value of 25% was employed for male and 35% for female to classify participants as “normal %BF” or “high %BF”. Thus, study participants were categorized as either “PWoH – normal %BF” (PWoH-NBF), “PWoH-high %BF” (PwoH-HBF), “PW –normal %BF” (PWH-NBF), or “PWH-high %BF” (PWH-HBF). Again, R package NanostringDiff was utilized for three pairwise DGE analyses of PWoH-HBF, PWH-NBF and PWH-HBF groups against a control group (PWoH-NBF), respectively. The three DGE analyses were adjusted for age, sex, race, ethnicity and batch.

Partial correlation analysis was carried out to understand the degree of linear association between each pair of phenotypic variables in our data. For this, we (i) compute the Spearman’s rank-order correlation coefficient for each pair of variables, while adjusting for the effects of sex, age, and race, using the pingouin package; (ii) generate a heatmap showing the correlation score for each pair where the color indicates the direction and intensity of the correlation; (iii) only annotate the score for correlations that show significance (t-test p-value below 0.05).

Multivariate regression analysis was performed to evaluate the effects of age, sex, race, ethnicity, %BF, and HIV status on hydroxyproline and endotrophin levels. For this, we (i) remove all records missing one or more of the set of required variables; (ii) cast each variable into their appropriate data types; (iii) apply a Box-Cox power transformation (iv) and run an Ordinary Least Squares (OLS) regression model using the stats package; (v) validate the model’s assumptions for linearity, non-normality of residuals/errors, and homoscedasticity. We then applied the fitted regression models to predict the change effect of HIV status on hydroxyproline and endotrophin, while adjusting for race, age, sex, and %BF. For this, we use the mean for each continuous covariate (age, %BF) and the mode for each categorical covariate (race, sex) to measure the effect of HIV status. These results are used to generate the adjusted bar plots (Supplemental figures 1 and 3) with the standard error bars shown. Pathway analysis was performed using gene set enrichment analysis (GSEA) with the FGSEA algorithm, implemented via the clusterProfiler and ReactomePA packages in the R programming language. First, each gene marker was assigned a score based on the fold-change and p-value from the differential gene expression analysis. The calculated scores were then used to create a ranked gene list, followed by mapping the gene identifiers to their corresponding ENTREZ IDs. The ranked list was then used to assess pathway enrichment across multiple databases (GO, KEGG, and REACTOME). Significantly enriched pathways were identified using a p-value cutoff (p < 0.1) after controlling for multiple hypothesis testing using false discovery rate adjustment.

### Data availability

Supporting data values of figures and tables are available as a supplemental Supporting Data Values file. Extra data are available from the corresponding author upon request. In addition, the NanoString raw can be accessed via FigShare (https://figshare.com/) at https://doi.org/10.6084/m9.figshare.28971413.v1

### Study Approval

All participants provided informed consent, and the study received approval from the University of California San Francisco (UCSF) Institutional Review Board (14–14128).

## Author Contributions

D.L.A., S.K.K., and P.W.H. conceptualized and designed the study. A.R., M.E., T.F., and J.G.-V. assisted with participant enrollment, coordinated research visits, collected biospecimens, and entered patient metadata into the study database. S.B.M., T.A.T.P., D.B., and D.L.A. processed blood and tissue samples and performed experiments. N.Z. and Z.A. developed the antibodies and the endotrophin ELISA assay. D.Bu. performed the endotrophin assay. M.K.C. and A.A. cleaned the data, performed analyses, and contributed to figure design, which was supervised by D.L.A. S.G.D. and P.E.S. provided resources and revised the manuscript for important intellectual content. D.L.A. and M.K.C. wrote the original draft. P.W.H., S.K.K and D.L.A. contributed to the review and editing of the manuscript. All authors reviewed and approved the final version of the manuscript.

## Acknowledgments

We are grateful to the SCOPE and IDEO study participants. We acknowledge current and former IDEO and SCOPE clinical study team members Melissa Buitrago, Fatima Ticas, Rebecca Hoh, Viva Tai, Yanel Hernandez, Sara Jara-Padilla. Koliwad laboratory members Abigail Steinmetz and Rachel Cheang. We acknowledge the contributions of the UCSF Clinical and Translational Science Unit, and AIDS Specimen Bank.

