## Supplemental data for "A Distinct Form of Subcutaneous Fat Fibrosis Predicts Insulin Resistance in People with HIV"

The PDF file includes Supplemental figures 1-5 and Supplemental Tables 1-5. Other Supplementary Material for this manuscript includes the following: Supporting Data Values file

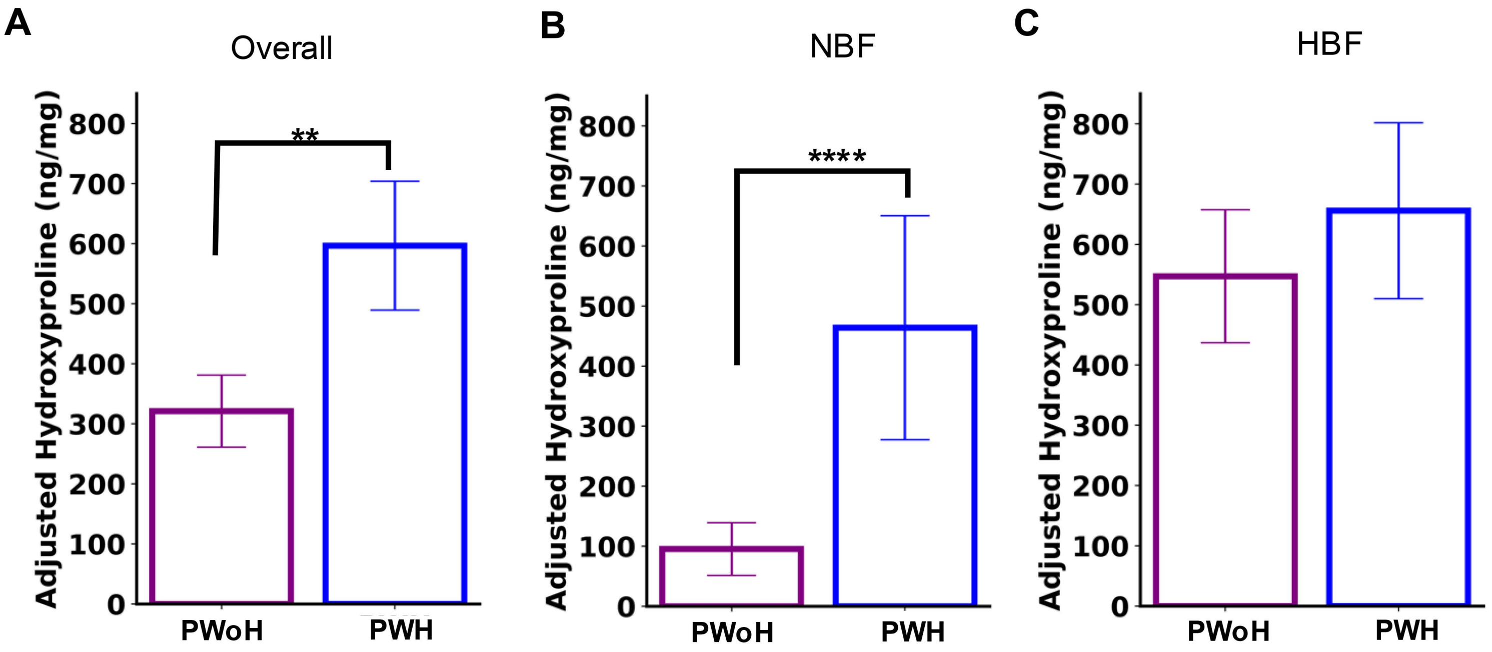


**Supplemental Figure 1.** Adjusted hydroxyproline (HYP) levels in the SAT of people with HIV (PWH) and people without HIV (PWoH), stratified by adiposity. HYP levels were compared between PWH and PWoH, adjusted for sex, age, and race/ethnicity, and further stratified into normal adiposity and high adiposity groups, as described in the methods. (A) PWH exhibited significantly higher SAT HYP levels vs. PWoH (PWoHn = 62, PWH n = 40, p 0.0038). (B) Subgroup analysis for participants with normal %BF (%BF < 25% for male, %BF < 35% for female) in the two groups (PWoH n = 29, PWH n = 17, p 0.0006) showing higher SAT HYP content in PWH-NBF (p <0.001). (C) Subgroup analysis for participants with high %BF (%BF ≥ 25% for male, %BF ≥ 35% for female) in the two groups (PWoH n = 33, PWH n = 23, p = 0.43) showing no difference in SAT HYP content between the two groups. Differences between the two groups were analyzed by using the model coefficient for HIV status, the p-value was computed from the t-statistic (the coefficient estimate divided by its standard error) using a t-distribution with appropriate degrees of freedom. ** <0.01, **** p < 0.001

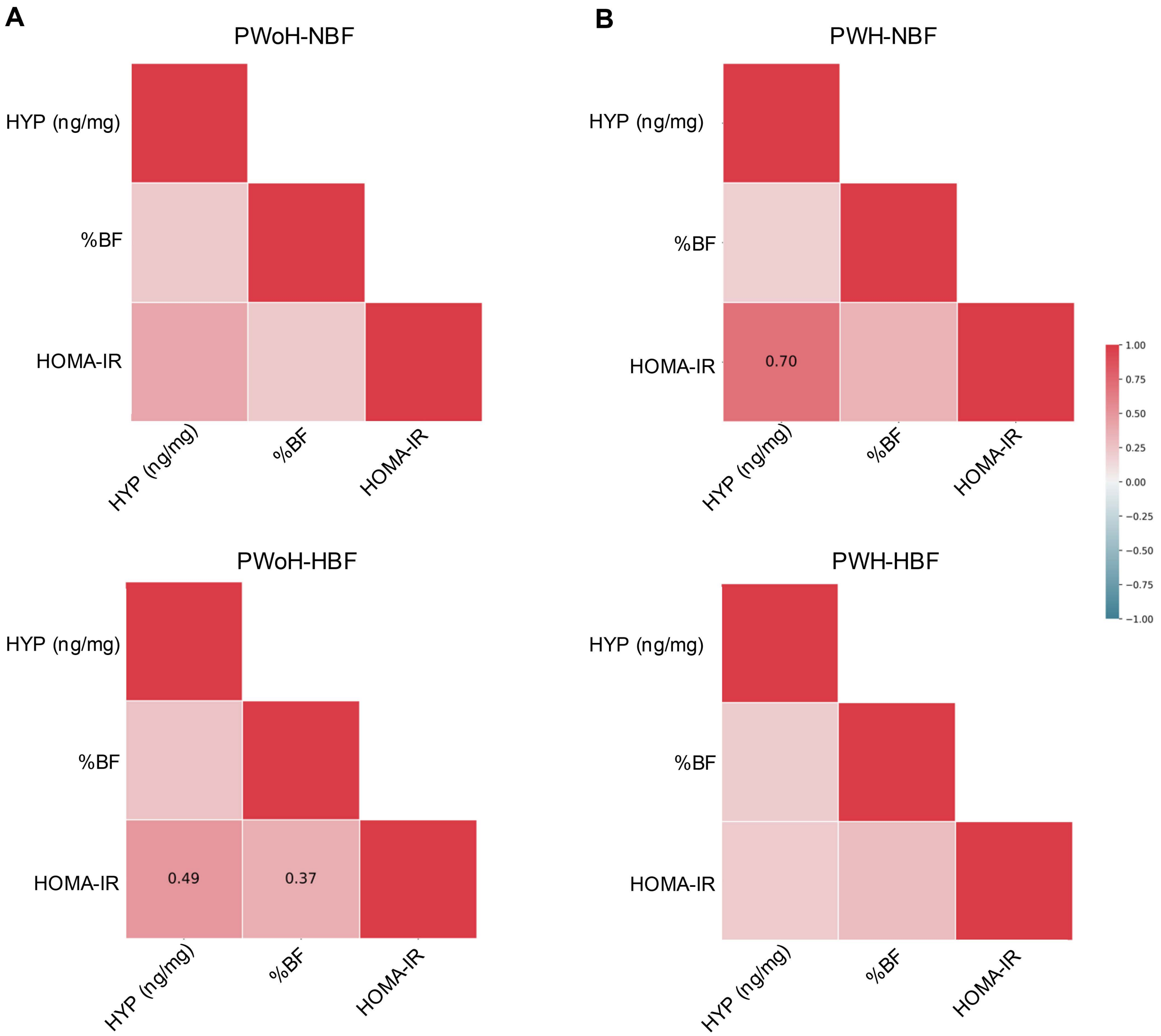


**Supplemental Figure 2.** Correlation analysis between SAT HYP levels, HOMA-IR, and % body fat in PWH and PWoH, stratified by normal and high adiposity. Spearman correlation analysis was performed to assess the relationships between hydroxyproline levels, HOMA-IR, and %BF in PWH and PWoH, stratified by NBF and HBF groups. (A) Correlation in PWoH-NBF (top panel, n= 29) and PWoH-HBF (lower panel, n= 33). (B) Correlation in PWH-NBF (top panel, n=17) and PWH-HBF (lower panel, n=23). Spearman's rank-order correlation coefficients were calculated for each pair of variables while adjusting for the effects of sex, age, and race. The partial Spearman's correlation coefficient rho is shown on the heatmap only for the correlations that meet statistical significance ($p < 0.05$). Significance levels (p-values) were computed from the partial correlation coefficients using a two-tailed t-test. HYP: Hydroxyproline

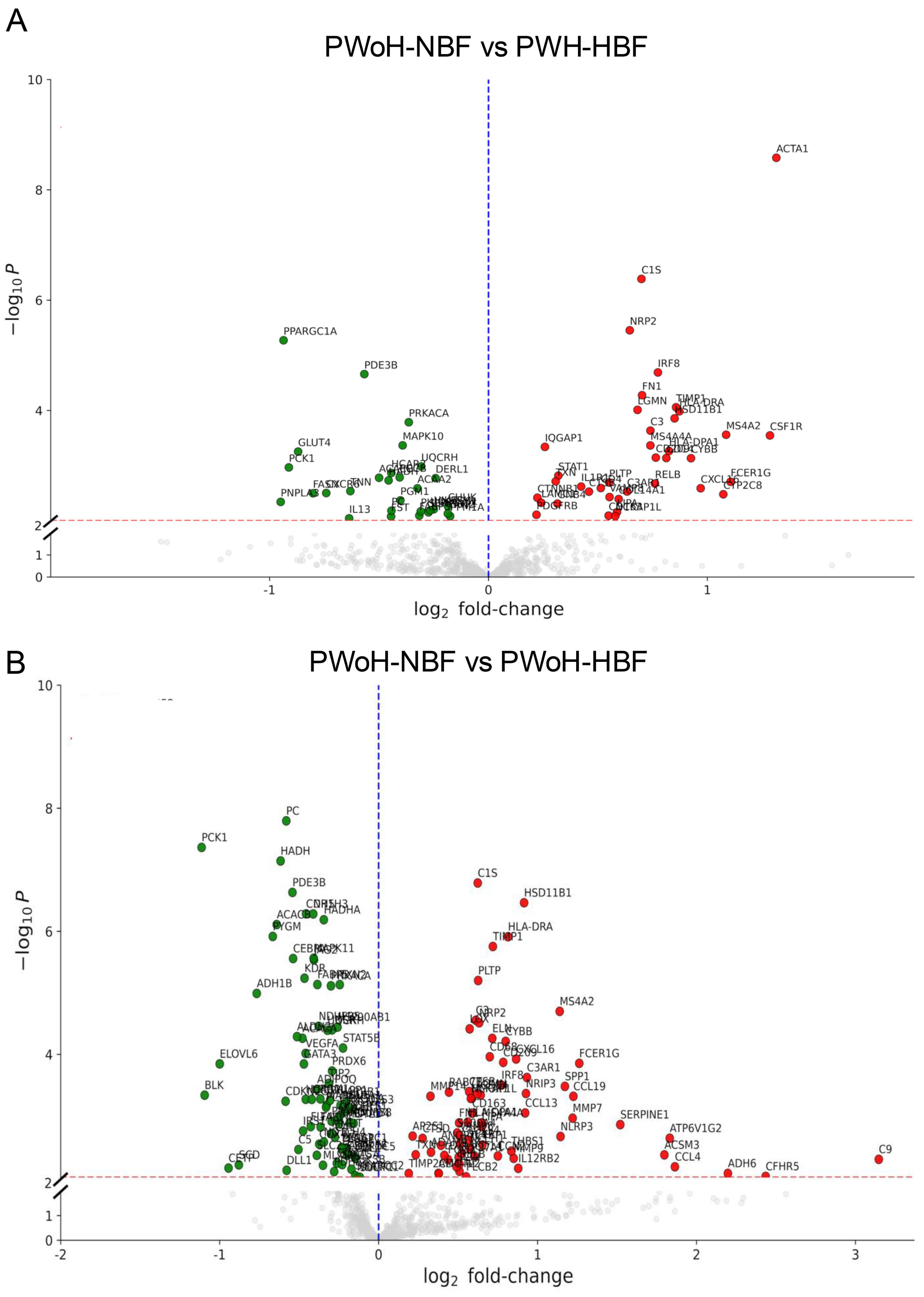


**Supplemental Figure 3.** Volcano plots showing differentially expressed genes (DEGs) in the SAT from (A) PWH-HBF (n = 21), and (B) PWoH-HBF (n = 20) each compared to the control group, PWoH-NBF (n = 18). DEGs were identified using linear modeling (limma), with a nominal p-value cutoff of 0.01 and $\log_2$ fold-change thresholds. Genes with increased expression in each comparison are shown in red and those with decreased expression in green.

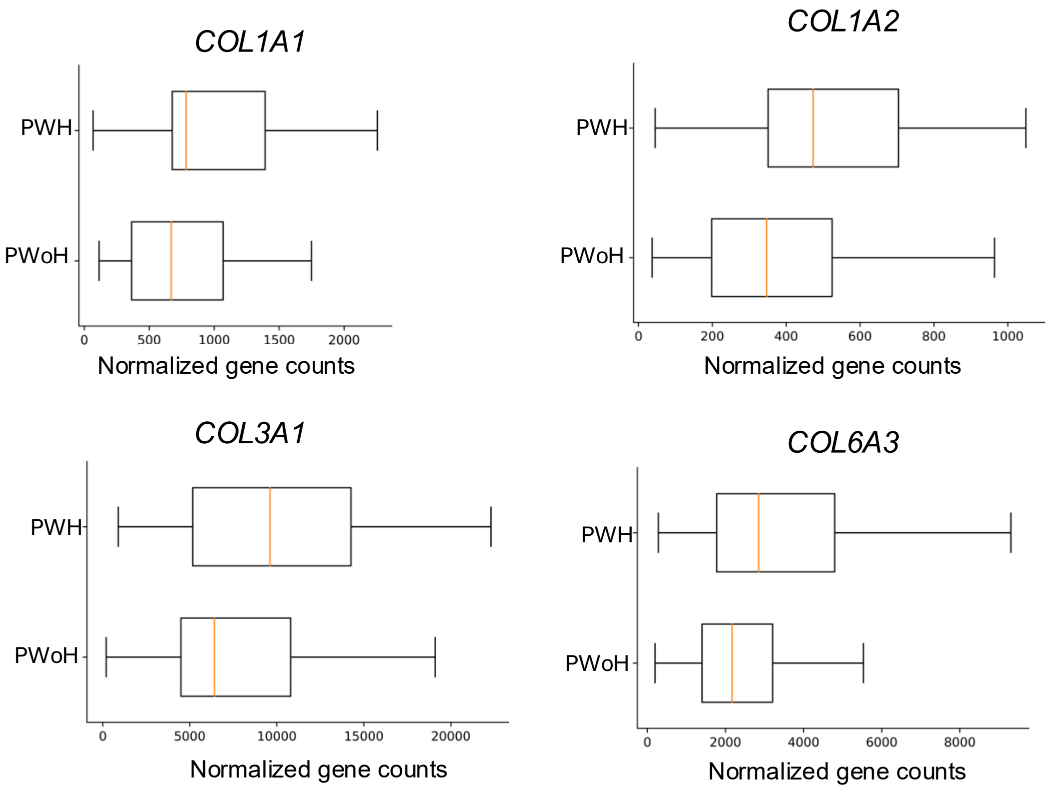


**Supplemental Figure 4.** Comparison of normalized expression of collagen-encoding transcripts in SAT from PWH and PWoH. NanoString-based quantification of *COL1A1*, *COL1A2*, *COL3A1*, and *COL6A3* expression in SAT samples from PWH and PWoH. Although differences between groups did not reach statistical significance ($p = 0.158$ for *COL1A1*; $p = 0.0548$ for *COL1A2*; $p = 0.131$ for *COL3A1*; $p = 0.0968$ for *COL6A3*), all transcripts showed a trend toward higher expression in PWH. All boxplot boxes display the interquartile range (IQR), with horizontal lines indicating the median. Statistical comparisons were performed using the Wilcoxon rank-sum test.

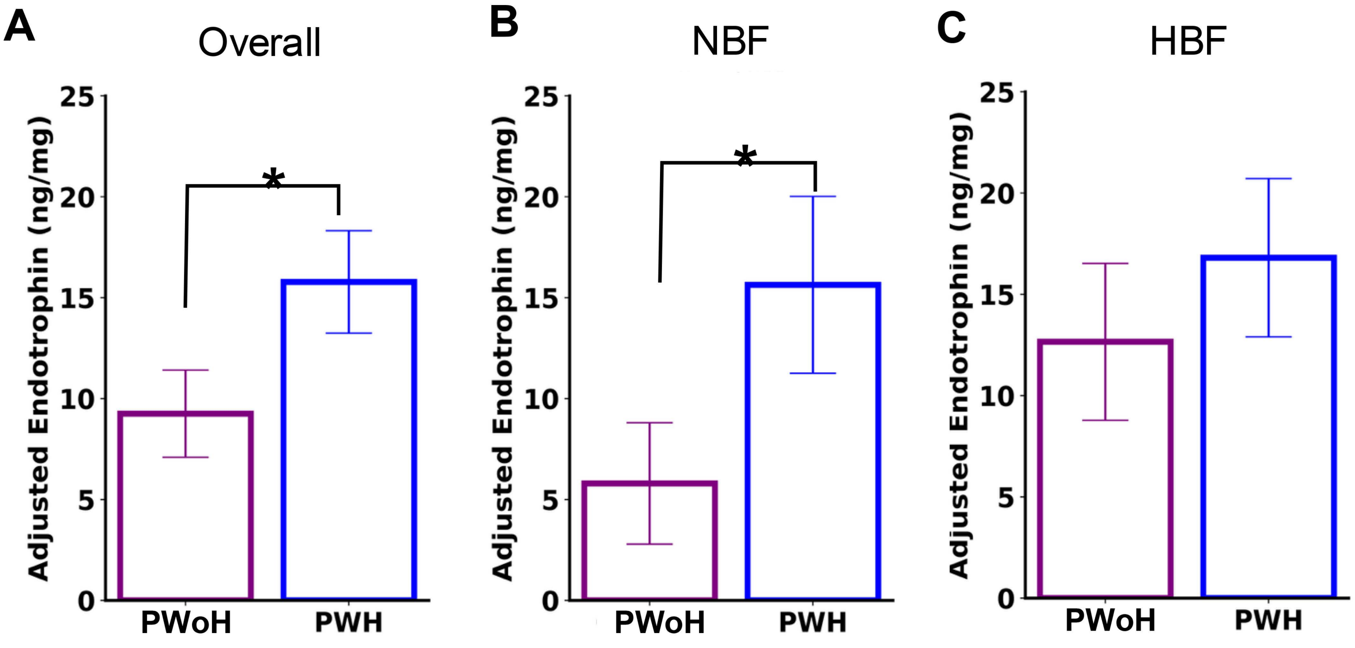


**Supplemental Figure 5**. Adjusted Endotrophin (ETP) levels in PWH and PWoH, stratified by adiposity. Endotrophin levels were compared between PWH and PWoH, adjusted for sex, age, and race/ethnicity, and further stratified into normal adiposity (NBF)and high adiposity groups (HBF), as described in the methods. (A) PWH exhibited significantly higher ETP levels compared to PWoH (PWoH n = 43, PWH n = 25, p 0.013). (B) Subgroup analysis for NBF participants in the two groups showed higher ETP levels in PWH-NBF vs. PWoH-NBF, (PWoH n = 19, PWH n = 9, p = 0.023). (C) Subgroup analysis for participants with HBF in the two groups showed no difference in ETP levels between the two groups (PWoH n = 24, PWH n = 16, p =0.30). Differences between the two groups were analyzed by using the model coefficient for HIV status, the p-value was computed from the t-statistic (the coefficient estimate divided by its standard error) using a t-distribution with appropriate degrees of freedom. * $p < 0.05$

**Supplemental Table 1**. Multivariable regression analysis was performed to evaluate the association between HIV status and VAT mass as a proportion of total body mass (%VAT), adjusting for sex, race/ethnicity, and age.

| Independent Variable | Coefficient | t-statistic | p-value |
|---|---|---|---|
| **Intercept** | 0.393 | 1.19 | 0.001 |
| **Sex (Male)** | 0.001 | 0.05 | 0.958 |
| **Race/Ethnicity** | | | |
| **Asian** | 0.027 | 0.248 | 0.632 |
| **Black** | 0.0759 | 0.203 | 0.805 |
| **Hispanic** | -0.004 | -0.004 | 0.968 |
| **More than one race** | 0.030 | 0.201 | 0.841 |
| **Native Hawaiian or Pacific Islander** | 0.0267 | 0.245 | 0.807 |
| **HIV Status** | 0.0239 | 0.884 | *0.379* |
| **Age** | 0.0636 | 5.833 | *0.000* |

The model aimed to determine whether the higher VAT levels observed in people with HIV (PWH) were attributable to HIV infection itself or other demographic and metabolic factors. Coefficients represent the direction and magnitude of the effect of each variable on %VAT. Statistically significant associations (< 0.05) are italicized. The model was fitted using data from 107 participants.

**Supplemental Table 2**. Results from a regression analysis examining the relationship between hydroxyproline levels in SAT and key independent variables, including HIV status, sex, race/ethnicity, age, and percentage body fat (%BF).

| Independent Variable | Coefficient | t-statistic | p-value |
|---|---|---|---|
| **Intercept** | -6.11 | -1.80 | 0.075 |
| **HIV status** | 3.79 | 2.96 | *0.004* |
| **Sex (Male)** | 6.50 | 4.75 | *0.0001* |
| **Race/Ethnicity** | | | |
| **Asian** | 2.55 | 2.07 | *0.0410* |
| **Black** | 0.717 | 0.466 | 0.642 |
| **Hispanic** | 2.02 | 1.23 | 0.221 |
| **More than one race** | 7.83 | 1.47 | 0.144 |
| **Native Hawaiian or Pacific Islander** | 5.91 | 1.15 | 0.253 |
| **Age** | -0.0117 | -0.284 | 0.777 |
| **%BF** | 0.513 | 6.37 | *0.0001* |

Coefficients represent the direction and magnitude of the effect of each variable on hydroxyproline levels. Statistically significant associations (< 0.05) are italicized. The model was fitted using data from 107 participants.

**Supplemental Table 3**. Characteristics of Study Participants included in SAT transcriptional analysis.

| | *PWoH (n=38)* | *PWH (n=37)* | *p-value* |
|---|---|---|---|
| **Age (years)** | 46 ± 13 | 53 ± 11 | 0.02 |
| **Sex at birth** | | | |
| Female | 18 | 10 | 0.11 |
| Male | 20 | 27 | |
| **Race/Ethnicity** | | | |
| White | 12 | 13 | |
| Hispanic | 10 | 5 | |
| Asian | 14 | 1 | <0.01 |
| Black | 2 | 16 | |
| Other* | 0 | 2* | |
| **BMI (kg/m$^2$)** | 28.3 ±7.1 | 29.7 ± 7.6 | 0.21 |
| **Body Fat Percentage (%)** | 31.4 ± 7.9 | 29.7 ± 7.8 | 0.28 |
| **VAT mass/total mass (%)** | 0.7 ± 0.3 | 0.7 ± 0.3 | 0.24 |
| **Android Gynoid Ratio** | 1.0 ± 0.2 | 1.1 ± 0.2 | 0.06 |
| **Fasting glucose (mg/dL)** | 93.0 (87.3, 100.8) | 91.0 (85.0, 101.0) | 0.40 |
| **Fasting insulin (mU/L)** | 12.3 (5.9, 17.1) | 10.0 (7.2, 16.7) | 0.95 |
| **HOMA-IR** | 2.9 (1.4, 4.3) | 2.7 (1.5, 3.8) | 0.81 |
| **Hydroxyproline (ng/mg fat)** | 188.5 (62.9, 665.5) | 570.9 (346.4, 899.6) | <0.01 |

Values are presented as mean ± SD or median (interquartile range). Differences between the groups were analyzed by Chi- Square test for categorical variables and Wilcoxon rank-sum test for continuous variables. *Other category for race and ethnicity includes one individual identifying Hawaiian or Pacific islander and another individual identifying as more than one race. VAT: Visceral adipose tissue by DEXA

**Supplemental Table 4**. Genes differentially expressed in adipose Tissue across groups compared to Control (PWoH-NBF).

### A. DGE analysis of PWH-NBF group vs control (PWoH-NBF)

| **Upregulated genes** | | |
|---|---|---|
| **Gene** | **logFC** | **p-value** |
| LIPG | 37.0635493962762 | 0.0001790040936917 |
| NCR1 | 19.068820852495 | 0.0 |
| CYP2C19 | 10.9065936743038 | 0.0018029211518294 |
| SPIB | 3.2161990953505 | 4.22782012320377e-05 |
| IL10 | 2.89608777943861 | 1.1478225037109302e-09 |
| MMRN1 | 2.62000199673392 | 1.05170915538633e-05 |
| SH2D1A | 2.16852664015033 | 0.0010360068272776 |
| CXCL8 | 2.14797295958307 | 0.0054244893486407 |
| APOC2 | 2.06269392078174 | 0.001046251873389 |
| PTGS2 | 1.87008454986986 | 0.0075826384495024 |
| GBP5 | 1.76619746503618 | 0.0010166964112662 |
| KLRK1 | 1.74862257605654 | 0.0020590839783849 |
| CCL4 | 1.72537703969768 | 0.0085974098141412 |
| RETN | 1.68515939979527 | 0.005618064691199 |
| AIM2 | 1.67229338015887 | 8.99083687693203e-05 |
| EGR1 | 1.59633666406913 | 0.0074415148706591 |
| NKG7 | 1.57013587748095 | 0.0039954064302706 |
| PRF1 | 1.51998468841396 | 0.0044384941338289 |
| NLRP3 | 1.50825597798173 | 1.41565934649313e-05 |
| CSF1R | 1.47868595986773 | 6.26688053280144e-05 |
| MS4A2 | 1.35122697661202 | 4.02261356069467e-06 |
| CYBB | 1.32549300458338 | 8.59910130879271e-08 |
| CXCL2 | 1.2860523601295 | 0.0042026028817497 |
| GZMA | 1.22598748179971 | 0.0031535967329403 |
| CYP2C8 | 1.20418780889042 | 0.0058212390514509 |
| DPP4 | 1.12778518462704 | 1.47801709993178e-06 |
| TIMP1 | 1.09537372981371 | 1.22880864551478e-05 |
| CXCR4 | 1.08484348837334 | 0.0060632998740936 |
| CXCL16 | 1.05575757718985 | 0.0002358358398981 |
| C3 | 0.987164430175923 | 3.47724303481067e-05 |
| MS4A4A | 0.880679247232368 | 0.0003666789341642 |
| DOCK2 | 0.85588163831463 | 0.0040055089239716 |
| MS4A1 | 0.853524533937161 | 0.0073948042816454 |
| NCKAP1L | 0.827969055524931 | 0.0003151521138139 |
| CD209 | 0.809845614209916 | 0.0017379738733492 |
| ARG1 | 0.787332742751883 | 1.59929821608173e-08 |
| VAMP8 | 0.755783596934542 | 0.0001617320904835 |
| IRF8 | 0.690191366475388 | 0.0085027642567265 |
| C1S | 0.64649005197319 | 2.73718970381642e-05 |
| COL14A1 | 0.614968575028804 | 0.0078796539775974 |

| | | |
|---|---|---|
| LEPR | 0.599595757415699 | 0.0033620671485361 |
| MAF | 0.477616137601984 | 0.0001633073830555 |
| DIAPH1 | 0.473089837835164 | 0.0048593626965918 |
| PELI1 | 0.462036621923058 | 0.0065846217918059 |
| MAP3K1 | 0.389395592798025 | 0.0057690823053355 |
| PRKACB | 0.288497049391855 | 0.0008565346200428 |
| GNPTAB | 0.284313930188901 | 0.0022743032662968 |

| **Downregulated genes** | | |
|---|---|---|
| **Gene** | **logFC** | **p-value** |
| BPI | -13.6350800120929 | 0.0012476501139232 |
| CETP | -2.0569002492951 | 3.33365395858154e-05 |
| PNPLA3 | -1.66806342285739 | 0.0001022432495472 |
| ELOVL6 | -1.6033781732963 | 0.0011618160826398 |
| TNN | -0.957132836888991 | 0.0003934214344714 |
| FASN | -0.948841872491082 | 0.0007606698618418 |
| GLUT4 | -0.83650608306932 | 0.0006789587624743 |
| SREBF1 | -0.676939092253767 | 0.0056576023422321 |
| NID2 | -0.640363751761175 | 0.000837357494457 |
| CDH5 | -0.629302385607305 | 0.0002315723072925 |
| PDE3B | -0.620047708618986 | 4.42804585722989e-05 |
| HSPG2 | -0.616283579833148 | 1.03686754193699e-06 |
| POSTN | -0.609877834688399 | 0.0023396135379374 |
| PC | -0.608695686721639 | 0.0004720351184858 |
| COX4I2 | -0.602271507186444 | 0.0059489582073862 |
| MAPK11 | -0.591904318443648 | 1.49007828742498e-07 |
| NR1H3 | -0.582230576520968 | 6.26717203688898e-08 |
| JAG2 | -0.56119435750372 | 1.83796849718698e-05 |
| MLXIPL | -0.540527923128575 | 0.000450669363187 |
| CEBPA | -0.53568268255011 | 0.002617773314925 |
| EPAS1 | -0.529158622151682 | 2.8874565122905e-05 |
| PDGFB | -0.521607111313353 | 0.0056309973008076 |
| FST | -0.5163404225087 | 0.0039254396218825 |
| SMAD6 | -0.502739071573549 | 0.0043387355176594 |
| COL4A2 | -0.498507151294085 | 0.000597836015315 |
| TEK | -0.487197807227465 | 0.0038616852085111 |
| MAP2K1 | -0.458760640960537 | 0.0067855143016536 |
| KDR | -0.458411407067871 | 0.0020342345446497 |
| PCCB | -0.443396439314582 | 0.0020181282468241 |
| RAPGEF2 | -0.430733182060567 | 0.0002296111593641 |
| ACACB | -0.419789251432895 | 0.0052272623713116 |
| PDE2A | -0.415391421122774 | 0.0070291129866049 |
| PRKACA | -0.407784528240615 | 0.0001182286470623 |
| CCL19 | -0.354869270739077 | 8.803086037900701e-10 |
| ALDH3A2 | -0.35255215463209 | 0.0028859438807122 |
| PDGFRB | -0.336911472734327 | 0.0007063147678153 |

| | | |
|---|---|---|
| STAT5A | -0.33318005449551 | 0.000784889496348 |
| SDC3 | -0.31365687531636 | 0.0026808984616407 |
| FNIP2 | -0.31006899657869 | 2.94657900329343e-05 |
| AMOTL1 | -0.294707319066912 | 0.001924882763945 |
| PLCG1 | -0.291556348514376 | 0.0054067983202873 |
| MAP1LC3A | -0.283025037078336 | 0.0025819512463636 |
| CSNK1E | -0.279775017064461 | 0.0001530476947231 |
| MTOR | -0.267093900668617 | 0.0052081204172618 |
| PTPN12 | -0.261336320038473 | 0.0048055500213048 |
| ABCB11 | -0.229503234711678 | 0.0045765764338345 |

**B. DGE analysis of PWoH-HBF group vs control (PWoH-NBF)**

| **Upregulated genes** | | |
|---|---|---|
| **Gene** | **logFC** | **p-value** |
| LIPG | 12.5658578489718 | 3.62341552695433e-06 |
| NCR1 | 6.53321364098701 | 0.0012942409603019 |
| RETN | 4.27954128574369 | 7.31762317407458e-10 |
| C9 | 3.14655026412957 | 0.0050694825078867 |
| CFHR5 | 2.4348348139527 | 0.0096030675966992 |
| ADH6 | 2.19726094093612 | 0.008583766316398 |
| CCL4 | 1.86393411228154 | 0.0066935171144165 |
| ATP6V1G2 | 1.83158357650667 | 0.002303985931377 |
| ACSM3 | 1.79788766280471 | 0.0042975103005348 |
| SERPINE1 | 1.52039642104054 | 0.0013841780915128 |
| FCER1G | 1.26215183108552 | 0.0001397818101193 |
| CCL19 | 1.22523387493903 | 0.0004799063603164 |
| MMP7 | 1.22066071550097 | 0.001087217963193 |
| SPP1 | 1.17181881193815 | 0.000331135891023 |
| NLRP3 | 1.14441998822852 | 0.0021583594765746 |
| MS4A2 | 1.13918199983023 | 2.00021014687968e-05 |
| C3AR1 | 0.932883313396239 | 0.0002374039383166 |
| NRIP3 | 0.926804309652363 | 0.0004330707091608 |
| CCL13 | 0.921876398438541 | 0.0008948767214692 |
| HSD11B1 | 0.915228122215726 | 3.43586869289148e-07 |
| IL12RB2 | 0.878383007574826 | 0.0071108155415906 |
| CXCL16 | 0.864995782004825 | 0.0001190916462278 |
| MMP9 | 0.84973132105323 | 0.00488025566264 |
| THBS1 | 0.834885439816972 | 0.0037274784970192 |
| HLA-DRA | 0.813948838280461 | 1.22476188668674e-06 |
| CYBB | 0.79928515600563 | 6.14189542713639e-05 |
| CD209 | 0.784001586260263 | 0.0001347420410335 |
| IRF8 | 0.773114350706196 | 0.0003153459784946 |
| CCN2 | 0.750328929493782 | 0.0044857899332194 |
| TIMP1 | 0.71885443056538 | 1.76476995616959e-06 |
| ELN | 0.71501203388943 | 5.48624285757748e-05 |
| CD68 | 0.699240941495151 | 0.0001093597968473 |

| | | |
|---|---|---|
| FBP1 | 0.65323585315868 | 0.00295606554928 |
| MS4A4A | 0.649339832713712 | 0.0012942283254676 |
| LIPA | 0.648319681561049 | 0.0014916344222107 |
| CD84 | 0.642307928068368 | 0.0004584306611667 |
| NRP2 | 0.632648454405621 | 3.06606317206359e-05 |
| PLTP | 0.626307453812017 | 6.3156214771265e-06 |
| C1S | 0.623583253832885 | 1.64045003137048e-07 |
| LGMN | 0.620682673651224 | 0.0004347302549648 |
| C3 | 0.611197931426978 | 2.78765264656311e-05 |
| CD14 | 0.602619810311084 | 0.0044572310742825 |
| CD163 | 0.588451072627671 | 0.0009049142397498 |
| HMOX1 | 0.586411378897562 | 0.0005199881809367 |
| NCKAP1L | 0.580106951612922 | 0.0005191275968708 |
| NCEH1 | 0.57360755102103 | 0.0033500894402736 |
| LOX | 0.572798588573352 | 3.84469098440032e-05 |
| CTSB | 0.569324147357392 | 0.0004019259470481 |
| LOXL4 | 0.56495195245715 | 0.0024676305124137 |
| HLA-DPA1 | 0.560751528378488 | 0.0012590552922965 |
| PLCB2 | 0.550457700937982 | 0.0097506033482832 |
| SYK | 0.524265076539803 | 0.0040097593294381 |
| ADCY7 | 0.510850594894949 | 0.0028461187619276 |
| RELB | 0.509027252397552 | 0.0060419168740232 |
| LEP | 0.508994076753643 | 0.007951821534137 |
| VAMP8 | 0.508783645342527 | 0.0021037745186167 |
| FN1 | 0.50621204841854 | 0.0012992565003806 |
| CYP27A1 | 0.503139320561746 | 0.0047084944490642 |
| S100A4 | 0.494569226803619 | 0.0018880113274336 |
| SCIN | 0.489986756643088 | 0.0067035546620628 |
| RAB7B | 0.442775376738002 | 0.0004100095535358 |
| FGD2 | 0.435975399479966 | 0.0051089515181564 |
| LOXL1 | 0.413586132703079 | 0.004373035232147 |
| ANGPTL4 | 0.392429176791241 | 0.0029974353395608 |
| CD4 | 0.378309098797065 | 0.008598962402476 |
| AMOTL2 | 0.376589253355955 | 0.0085686395049057 |
| ADAM9 | 0.329452296086357 | 0.003881006366963 |
| MMP14 | 0.326174199686071 | 0.0004782246489803 |
| CTSD | 0.276603880123539 | 0.0023192573621582 |
| TXN | 0.233127936876404 | 0.0042759057413326 |
| AP2S1 | 0.21372212042885 | 0.002126403997457 |
| TIMP2 | 0.189604114167868 | 0.0086154154113599 |

| Downregulated genes | | |
|---|---|---|
| **Gene** | **logFC** | **p-value** |
| GLUT4 | -1.40101915717033 | 9.76996261670138e-15 |
| FASN | -1.18275029701123 | 6.917155737085069e-11 |
| PCK1 | -1.1135855375738 | 4.3349399736492e-08 |
| BLK | -1.09477343005731 | 0.0004592893902254 |
| ELOVL6 | -0.999807176935802 | 0.0001425600249209 |
| CETP | -0.944404700876005 | 0.0070354767685487 |
| SCD | -0.879282150844023 | 0.0062403248728577 |
| ADH1B | -0.766834889001678 | 1.01989999554686e-05 |
| PYGM | -0.666109008384097 | 1.21306482170347e-06 |
| ACACB | -0.641033901732863 | 7.76917512657072e-07 |
| HADH | -0.61682695699948 | 7.18609588501451e-08 |
| CDKN2C | -0.585374702146896 | 0.0005767467215603 |
| PC | -0.580985233364473 | 1.60479999289365e-08 |
| DLL1 | -0.578634544880254 | 0.007626039603297 |
| PDE3B | -0.542265071563255 | 2.33455729747867e-07 |
| CEBPA | -0.538063175912794 | 2.77019753724961e-06 |
| ALDH2 | -0.513400726443074 | 5.13991811166292e-05 |
| C5 | -0.504605879705093 | 0.0035130147150455 |
| ACACA | -0.479101377901258 | 5.46839589853354e-05 |
| IRS1 | -0.475238387609918 | 0.0017552187246054 |
| GATA3 | -0.470176033894535 | 0.0001429802304749 |
| KDR | -0.466403895807412 | 5.78243006421619e-06 |
| VEGFA | -0.459396552104408 | 9.63067013639574e-05 |
| HCAR2 | -0.458480860612421 | 0.0005366876772771 |
| CDH5 | -0.456636746580115 | 5.20040980611647e-07 |
| FLT4 | -0.42735375194404 | 0.0014896635216823 |
| LPL | -0.420830664296778 | 0.0005383907436656 |
| NR1H3 | -0.411414773212982 | 5.22038172023898e-07 |
| MAPK11 | -0.408637050444612 | 2.73361417268969e-06 |
| JAG2 | -0.406667343595513 | 2.9045662952587e-06 |
| SLC25A10 | -0.388730516833104 | 0.0043644555000567 |
| ADIPOQ | -0.387448235270593 | 0.0003685867953167 |
| FABP5 | -0.385256766632489 | 7.31036803192087e-06 |
| NDUFB5 | -0.377114815401779 | 3.48664563948331e-05 |
| COX4I2 | -0.373819204435599 | 0.002874360124966 |
| PGM1 | -0.366802719788091 | 0.0005281797472608 |
| FABP4 | -0.365184098734083 | 0.0015120618095565 |
| MLXIPL | -0.350034571826249 | 0.0063326944364364 |
| HADHA | -0.344775680494745 | 6.49079894166071e-07 |
| NOTCH4 | -0.344320605008167 | 0.0026522399243397 |
| PCCB | -0.331047752402162 | 0.0007227820447424 |
| MAPK10 | -0.327675765657396 | 0.0006704854650951 |
| UQCRH | -0.321864053908682 | 4.05755232337768e-05 |
| TJP2 | -0.312797393493164 | 0.0002954065575719 |
| PHLPP1 | -0.305801403417391 | 0.0005563167764883 |
| PRKACA | -0.30027506787249 | 7.6858501332211e-06 |

| | | |
|---|---|---|
| PPARG | -0.295793471070289 | 0.0012177328659072 |
| MMUT | -0.295186983898824 | 0.001917434852404 |
| INSR | -0.293530214319915 | 3.99948317140186e-05 |
| PRDX6 | -0.292580822502326 | 0.0001863440624997 |
| PDHA1 | -0.279882315239398 | 0.0080292576866735 |
| CAT | -0.275013526568083 | 0.0022235756775518 |
| COX7C | -0.26308084309273 | 0.0058624226438863 |
| HSP90AB1 | -0.25892121218838 | 3.59923762460745e-05 |
| COX6B1 | -0.25149705958924 | 0.0010461489601038 |
| AMOTL1 | -0.248496458500018 | 0.0013404548003226 |
| TXN2 | -0.244769550094604 | 7.34450642925211e-06 |
| UQCRFS1 | -0.244032457061796 | 0.0009874789872911 |
| NDUFA1 | -0.239614728724516 | 0.0006698386209393 |
| EPAS1 | -0.232397609693857 | 0.0032441752398965 |
| PLCG1 | -0.229770775224642 | 0.003403874784404 |
| STAT5A | -0.229159918484354 | 0.0061862676975333 |
| STAT5B | -0.224256442047787 | 7.87086188998032e-05 |
| TBC1D4 | -0.2145071328696 | 0.0008537492952395 |
| ARRB1 | -0.203679268564543 | 0.0005897949331725 |
| ANAPC1 | -0.200186470682122 | 0.0033039405932952 |
| CSNK1E | -0.199390615620359 | 0.0043737030908596 |
| COX6A1 | -0.19775080600755 | 0.0040312001686508 |
| PIK3CA | -0.189354098806038 | 0.0007465230171317 |
| NDUFB8 | -0.179189419599557 | 0.0012761341527126 |
| GSK3B | -0.170883957908333 | 0.007368341502714 |
| NDUFS3 | -0.161893082923002 | 0.0007849462597843 |
| SMAD4 | -0.1573222844699 | 0.0013208071393937 |
| DEPDC5 | -0.152432825170134 | 0.004555761478603 |
| CUL1 | -0.147471440326502 | 0.0048231329889482 |
| SMARCC2 | -0.146904294331555 | 0.0093182566330254 |
| NDUFC1 | -0.135439547706931 | 0.0098393819186284 |
| SUGT1 | -0.118470252492207 | 0.0099051922926212 |

**C. DGE analysis of PWH-HBF group vs control (PWoH-NBF)**

| Upregulated genes | | |
|---|---|---|
| **Gene** | **logFC** | **p-value** |
| LIPG | 51.7934541624432 | 0.0007670645787504 |
| ACTA1 | 1.31521579566089 | 2.6199127312054303e-09 |
| CSF1R | 1.28607706119736 | 0.0002836045822766 |
| FCER1G | 1.10615102673695 | 0.0019660664287721 |
| MS4A2 | 1.08577814899163 | 0.0002756435982884 |
| CYP2C8 | 1.07298775304198 | 0.0033107764128944 |
| CXCL16 | 0.969499237797545 | 0.0025711882361325 |
| CYBB | 0.925214630034224 | 0.0007330596655099 |
| HLA-DRA | 0.871776762190134 | 0.0001034744414956 |
| TIMP1 | 0.857087878175498 | 8.75360201570974e-05 |
| HSD11B1 | 0.850153876669095 | 0.0001390562300681 |
| HLA-DPA1 | 0.82484930682703 | 0.0005430966531789 |
| CD14 | 0.812499792863197 | 0.0007282987166237 |
| IRF8 | 0.774056971637754 | 2.04200191141757e-05 |
| CD209 | 0.764464982374589 | 0.0007158957145183 |
| RELB | 0.76040063883743 | 0.0021126618067225 |
| C3 | 0.740007225545601 | 0.0002323098574248 |
| MS4A4A | 0.7396913849466 | 0.0004311018261131 |
| FN1 | 0.701877692410896 | 5.29213589371968e-05 |
| C1S | 0.698939403803965 | 4.11953550827349e-07 |
| LGMN | 0.680979205930875 | 9.73810544181442e-05 |
| NRP2 | 0.645633619209431 | 3.51276876076057e-06 |
| C3AR1 | 0.633208248075388 | 0.0029529358703718 |
| COL14A1 | 0.594842172597955 | 0.0040452371419992 |
| LIPA | 0.587397220409585 | 0.0069749489382144 |
| NCKAP1L | 0.579366975920453 | 0.0081984968167679 |
| VAMP8 | 0.553041926410598 | 0.0037092161975081 |
| PLTP | 0.551415878283834 | 0.0020246025205762 |
| CD163 | 0.549061108234821 | 0.0080682978277488 |
| CD4 | 0.513230166291694 | 0.0025475881952558 |
| CTSB | 0.459042808274346 | 0.0029627473184593 |
| IL1R1 | 0.423441568637876 | 0.0023967053946589 |
| STAT1 | 0.319108011343229 | 0.0015196091224339 |
| GNB4 | 0.315305240434932 | 0.0048763080679163 |
| TXN | 0.307092337829281 | 0.0019305605783678 |
| IQGAP1 | 0.258051334944683 | 0.000457322531995 |
| LAMC1 | 0.241128204887299 | 0.0047149751163261 |
| CTNNB1 | 0.223568462736151 | 0.0038174897512947 |
| PDGFRB | 0.219260195058869 | 0.0077722500957997 |

| Downregulated genes | | |
| --- | --- | --- |
| **Gene** | **logFC** | **p-value** |
| PNPLA3 | -0.949412949739135 | 0.0045547719061045 |
| PPARGC1A | -0.936392248221363 | 5.34019315068246e-06 |
| PCK1 | -0.911969703267581 | 0.0010765605939181 |
| GLUT4 | -0.869701342810029 | 0.0005560243594743 |
| FASN | -0.80171186783487 | 0.0031724367735293 |
| CXCR6 | -0.74108032987111 | 0.0031435459535992 |
| IL13 | -0.636335606472764 | 0.0090377834385504 |
| TNN | -0.631138216401729 | 0.0028948888127891 |
| PDE3B | -0.567215381335517 | 2.1915825292651e-05 |
| ACACB | -0.499906665023648 | 0.0016587497563203 |
| HADH | -0.456166894042748 | 0.0018512353297931 |
| FST | -0.445010658112409 | 0.0083524180243366 |
| HCAR2 | -0.443912413520443 | 0.0013581130585261 |
| PC | -0.443258169025953 | 0.0065701830402974 |
| PCCB | -0.405703602726079 | 0.0016325406722892 |
| PGM1 | -0.402472672959511 | 0.0042677386687005 |
| MAPK10 | -0.392024044215919 | 0.0004286712163288 |
| PRKACA | -0.364084979106198 | 0.0001633605283759 |
| ACAA2 | -0.324309659815205 | 0.0025750986715552 |
| FABP5 | -0.3158387764142 | 0.0080551403678366 |
| PHLPP1 | -0.309649497831601 | 0.0067658022283621 |
| UQCRH | -0.307050594221679 | 0.0010337588337037 |
| SERPINF1 | -0.273919401925337 | 0.0070465900594489 |
| UQCRFS1 | -0.265939596037728 | 0.0064626668777332 |
| HIKESHI | -0.250048062950452 | 0.0061906866429212 |
| DERL1 | -0.241601454505362 | 0.0016910847364358 |
| TXN2 | -0.185644457724275 | 0.0074213086399708 |
| CHUK | -0.184850748143104 | 0.0054636630601976 |
| PPM1A | -0.17513112581767 | 0.008262586733799 |

Log2(fold change) and p-value of differentially expressed genes in the three groups (PWoH-HBF (n=20), PWH-NBF (n=16), and PWH-HBF (n=21)), each compared to the control group (PWoH-NBF, n=18). (A) PWH-NBF compared to the Control-NBF group. 47 genes were upregulated, and 46 genes were downregulated. (B) Control-HBF compared to the Control-NBF group. 72 genes were upregulated, and 78 genes were downregulated. (C) PWH-HBF compared to the Control-NBF group. 39 genes were upregulated, and 29 genes were downregulated. All DGE analysis performed with NanostringDiff package on R. Statistical significance determined by p-value < 0.01.

**Supplemental Table 5**. Results from a regression analysis examining the relationship between Endotrophin (ETP) levels in plasma and key independent variables, including HIV status, sex, race/ ethnicity, age, and percentage body fat (%BF).

| Independent Variable | Coefficient | t-statistic | p-value |
|---|---|---|---|
| **Intercept** | 1.40 | 1.19 | 0.237 |
| **Sex (Male)** | 0.280 | 0.618 | 0.539 |
| **Race/Ethnicity** | | | |
| **Asian** | -0.260 | -0.481 | 0.632 |
| **Black** | 0.0759 | 0.203 | 0.840 |
| **Hispanic** | -0.332 | -0.808 | 0.422 |
| **More than one race** | -0.916 | -0.685 | 0.496 |
| **Native Hawaiian or Pacific Islander** | -0.335 | -0.269 | 0.789 |
| **HIV Status** | 0.901 | 2.57 | *0.013* |
| **Age** | 0.0086 | 0.689 | 0.493 |
| **%BF** | 0.0335 | 1.20 | 0.235 |

Coefficients represent the direction and magnitude of the effect of each variable on ETP levels. Statistically significant associations (< 0.05) are italicized. The model was fitted using data from 77 participants.
